# Glargine-to-intravenous insulin dose ratio, rebound hyperglycemia and hypoglycemia after discontinuation of insulin infusion in critically ill adults: a retrospective cohort study

**DOI:** 10.64898/2026.09.28.26364145

**Authors:** Lujun Shao, Yue Li, Li Wang, Wenbin Sun

## Abstract

**Background:** When intravenous insulin is stopped in critically ill patients, the basal insulin dose that balances rebound hyperglycemia against hypoglycemia is uncertain, because randomized trials were small and not designed to quantify this trade-off. We examined how the first glargine dose, relative to the preceding intravenous insulin requirement, was associated with both outcomes.

**Methods:** In this retrospective cohort study of the Medical Information Mart for Intensive Care IV database (2008–2022), we included transitions from intravenous insulin infusions of at least 6 hours to subcutaneous glargine in adults without diabetic ketoacidosis or hyperosmolar hyperglycemic state. The exposure was the ratio of the first glargine dose to 24 times the mean infusion rate over the final 6 hours. The primary outcomes, rebound hyperglycemia (glucose >180 mg/dL within 24 hours) and hypoglycemia (glucose <70 mg/dL within 48 hours) during intensive care unit follow-up, were analyzed with multivariable Cox models after multiple imputation.

**Results:** Among 6,220 transitions in 5,924 patients (87.5% after cardiac surgery), the median ratio was 0.41 (interquartile range 0.30–0.46). Kaplan–Meier cumulative incidences were 44.4% for rebound hyperglycemia (2,109 events) and 6.4% for hypoglycemia (211 events). Each 0.1 increase in the ratio was associated with less rebound hyperglycemia (hazard ratio 0.96, 95% confidence interval 0.93–0.99) and more hypoglycemia (1.11, 1.04–1.18), consistently across 21 sensitivity analyses. Rebound within the first 6 hours (57.6% of events) was not associated with the ratio (0.99, 0.95–1.02), whereas later rebound was (0.91, 0.86–0.96). Standardized to the cohort, a ratio of 0.60 versus 0.40 corresponded to 2.7 (0.6–4.9) fewer rebound and 1.7 (−0.2 to 3.5) more hypoglycemia episodes per 100 transitions. In secondary analyses, rebound was less frequent with glargine given at least 2 hours before discontinuation (0.86, 0.77–0.96) and more frequent with an oral diet (1.49, 1.30–1.71).

**Conclusions:** In critically ill adults, mostly after cardiac surgery, higher glargine doses relative to the preceding intravenous requirement were associated with less rebound hyperglycemia after the first 6 hours and more hypoglycemia, a gradual trade-off. Which basal fraction is preferable depends on the weight given to each outcome; a randomized trial including the guideline-implied fraction of about 0.30 is warranted.

## Background

Hyperglycemia is common in critically ill adults and is particularly frequent after cardiac surgery, where postoperative glucose values of 180 mg/dL or higher have been associated with more infections, longer hospital stays and higher costs in patients without diabetes [1, 2]. Continuous intravenous (IV) insulin is the preferred treatment for hyperglycemia in the intensive care unit (ICU) [3, 4]. Hypoglycemia is the main hazard of insulin therapy in this setting: both moderate and severe hypoglycemia have been associated with higher mortality in critically ill patients [5, 6].

Once patients stabilize, IV insulin must be converted to a subcutaneous regimen. This transition is a period of particular risk. The short half-life of IV insulin allows glucose to rise within hours after the infusion is stopped, whereas subcutaneous basal insulin takes several hours to act; conversely, insulin requirements often fall as the acute stress response resolves, so a basal dose calculated from the preceding infusion may become excessive [1, 7, 8]. The American Diabetes Association recommends a transition protocol with subcutaneous basal insulin given 2 h before the infusion is stopped and, for initial subcutaneous dosing in hospitalized patients, suggests using about 60% of the total daily dose estimated from the infusion rate, half of it as basal insulin, which implies a basal fraction of about 0.30 [4]. How rebound hyperglycemia and hypoglycemia change across the basal fractions used in practice, however, has not been quantified.

The evidence on this fraction is limited and inconsistent. Three small randomized trials of 75, 223 and 82 patients compared basal doses of 40% to 80% of the IV insulin requirement and did not identify a clearly superior dose [9–11]. Among observational studies, one found the highest proportion of glucose values in range when the initial basal dose was 50–59% of the IV requirement estimated from the final 6 h [12], and another in a medical ICU examined the timing rather than the size of the first basal dose [13]. These studies relied mainly on the proportion of glucose values in range, were underpowered for hypoglycemia, and did not describe how rebound hyperglycemia and hypoglycemia change across the range of doses used in practice or how this relationship depends on the timing of the first dose and on nutritional status.

We therefore used a large ICU database with time-stamped insulin infusion, medication administration and glucose records [14] to examine the association between the first glargine dose, expressed as a fraction of the preceding IV insulin requirement, and rebound hyperglycemia and hypoglycemia after discontinuation of IV insulin, and to assess the roles of the timing of glargine and of nutritional status.

## Methods

### Study design and data source

This retrospective cohort study used the Medical Information Mart for Intensive Care IV (MIMIC-IV), version 3.1, a de-identified database of patients admitted to the emergency department and ICUs of Beth Israel Deaconess Medical Center (Boston, MA, USA) between 2008 and 2022 [14–16]. Information on medications used before admission was obtained from the linked MIMIC-IV-Note (version 2.2) and MIMIC-IV-ED (version 2.2) modules [17, 18]. Data were extracted with Structured Query Language (SQL) on Google BigQuery (Additional file 1: Supplementary Methods). The study is reported in accordance with the Strengthening the Reporting of Observational Studies in Epidemiology (STROBE) and REporting of studies Conducted using Observational Routinely-collected health Data (RECORD) statements [19, 20]; the completed RECORD checklist, which incorporates the STROBE items, is provided in Additional file 2.

### Study population

The unit of analysis was the transition from an IV insulin infusion to subcutaneous glargine. Charted infusions of IV regular insulin with a positive rate were concatenated into infusion episodes; interruptions of 6 h or less were treated as part of the same episode, and episodes lasting at least 6 h were retained. The end of an episode defined discontinuation and time zero. We excluded episodes that occurred during an admission with a diagnosis of diabetic ketoacidosis or hyperosmolar hyperglycemic state, episodes without ICU observation after discontinuation, episodes without basal insulin from 12 h before to 1 h after discontinuation, episodes in which the first basal insulin in this window was not glargine, and episodes in which the exposure could not be calculated or exceeded 3 (Fig. 1). A patient could contribute more than one episode. Item identifiers and diagnosis and procedure codes are listed in Additional file 1: Table S1.

**Fig. 1.**
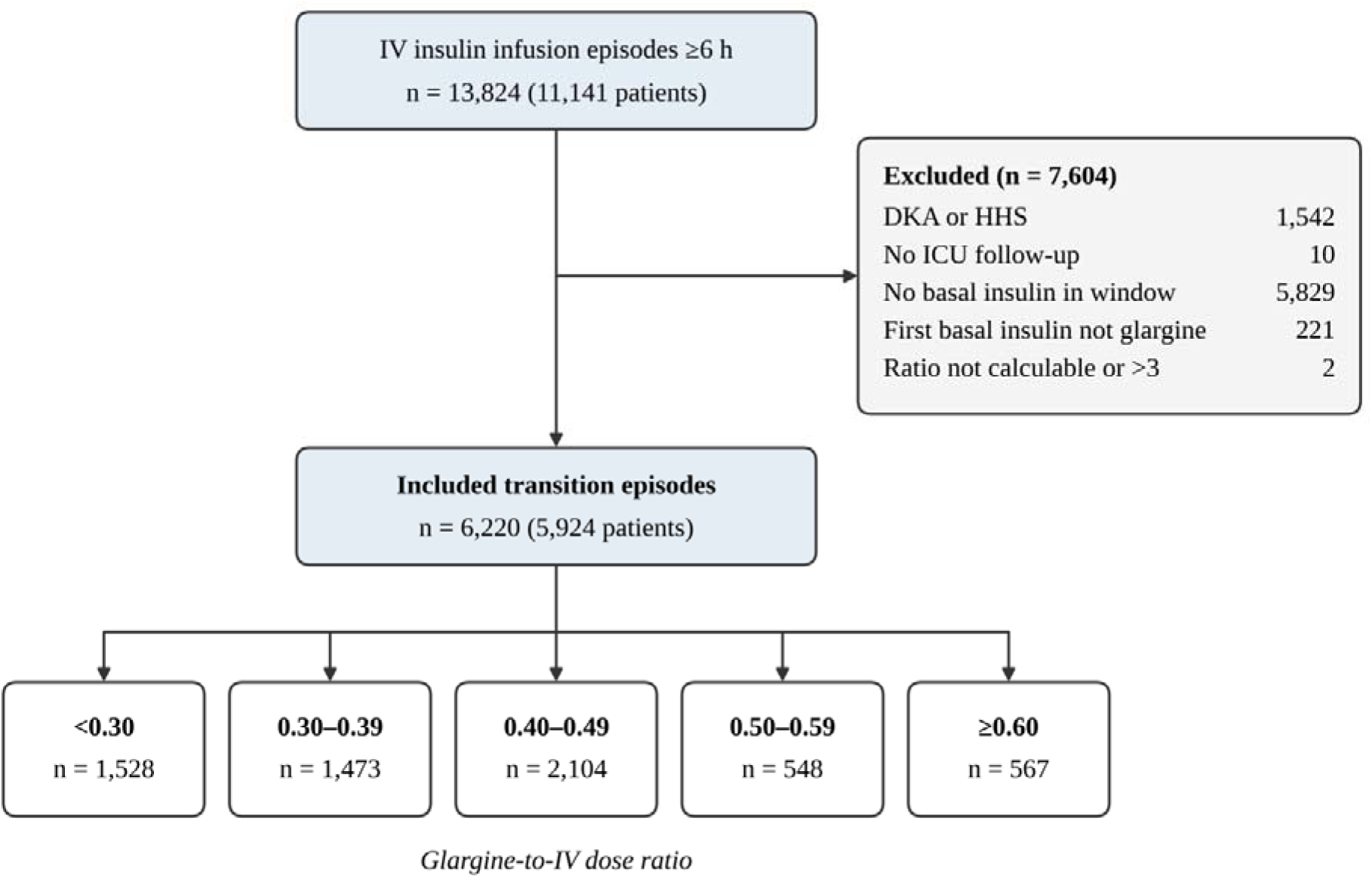
Selection of transition episodes from intravenous insulin to subcutaneous glargine. Infusion episodes comprised records of intravenous regular insulin with interruptions of 6 h or less and a total duration of at least 6 h. Exclusions were applied in the order listed. The exposure window extended from 12 h before to 1 h after discontinuation of the infusion. The ratio is the first glargine dose divided by 24 times the mean infusion rate over the final 6 h. DKA, diabetic ketoacidosis; HHS, hyperosmolar hyperglycemic state; ICU, intensive care unit; IV, intravenous.

### Exposure

The exposure was the glargine-to-IV dose ratio, defined as the first glargine dose divided by the estimated IV total daily dose. The IV total daily dose was calculated as the mean infusion rate over the final 6 h before discontinuation multiplied by 24, the approach used in the randomized comparison of glargine conversion doses by Schmeltz et al. [9]. The ratio was analyzed as a continuous variable (per 0.1 increase), with restricted cubic splines, and in five categories (<0.30, 0.30–0.39, 0.40–0.49, 0.50–0.59 and ≥0.60). The 0.40–0.49 category was the reference because it was the most frequently used and lies at the lower end of the range of basal fractions (0.40–0.80) evaluated in randomized trials [9–11].

Glargine administrations were identified in ICU charting and in the electronic medication administration record (eMAR); when no basal insulin was charted in the ICU record during the exposure window, the eMAR administration was used (Additional file 1: Table S2).

### Outcomes

The two primary outcomes were rebound hyperglycemia, defined as the first glucose value above 180 mg/dL within 24 h after discontinuation, and hypoglycemia, defined as the first glucose value below 70 mg/dL within 48 h. Glucose values comprised fingerstick, whole-blood and serum measurements charted in the ICU; values of 10 mg/dL or less or 1,000 mg/dL or more were removed, and the lowest value was used when several were charted at the same time. Secondary outcomes were severe hyperglycemia (above 250 mg/dL) within 24 h, clinically significant hypoglycemia (below 54 mg/dL) within 48 h [21], and resumption of IV insulin within 48 h.

Follow-up started at discontinuation and ended at the outcome, the end of the outcome window, ICU discharge or resumption of IV insulin, whichever occurred first; follow-up was restricted to the ICU (Additional file 1: Supplementary Methods). Glycemic control and insulin use after discontinuation were described.

### Covariates

Covariates were selected a priori as factors likely to influence both the glargine dose chosen and the glycemic outcomes. Patient characteristics were age, sex, diabetes (diagnosis codes), glycated hemoglobin (HbA1c), use of insulin before admission, body weight, serum creatinine, Charlson comorbidity index [22, 23] and Sequential Organ Failure Assessment (SOFA) score on the first ICU day [24, 25]. Characteristics of the infusion were its duration, the mean infusion rate over the final 6 h, the trend in insulin requirement (ratio of the mean rate in the final 6 h to that in the preceding 6 h), the final infusion rate, mean glucose over the final 6 h and the last point-of-care glucose. Treatment and other characteristics were vasopressor dose in norepinephrine equivalents [26], enteral or parenteral nutrition, an oral diet at discontinuation, dextrose infusion within the preceding 2 h, corticosteroid therapy, admission to the cardiac vascular ICU, cardiac surgery (coronary artery bypass grafting or valve surgery) before discontinuation, time in the ICU before discontinuation, basal insulin 12–48 h before discontinuation, glargine administration at least 2 h before discontinuation, and calendar period (anchor year group). Covariate definitions, time windows and missingness are given in Additional file 1: Table S3.

### Statistical analysis

Continuous variables are presented as medians (interquartile range [IQR]) and categorical variables as numbers (percentages). Cumulative incidence at 24 h (rebound and severe hyperglycemia) and at 48 h (hypoglycemia outcomes and resumption of IV insulin) was estimated with the Kaplan–Meier method; with censoring at ICU discharge, these estimates refer to risks under continued ICU observation. Associations between the ratio and each outcome were estimated as hazard ratios (HRs) with 95% confidence intervals (CIs) from Cox proportional hazards models with robust variance clustered by patient [27], fitted without and with adjustment for all covariates. Continuous covariates were modeled with restricted cubic splines with three knots at the 10th, 50th and 90th percentiles (vasopressor dose, which was zero in most episodes, was modeled linearly) to avoid residual confounding from misspecified functional forms, which is of particular concern because the exposure is mathematically coupled to the infusion rate [28]. The dose–response relationship was modeled with restricted cubic splines with four knots at the 5th, 35th, 65th and 95th percentiles [28], with 0.40 as the reference, and nonlinearity was tested with a Wald test of the nonlinear terms. The proportional hazards assumption was assessed with scaled Schoenfeld residuals [29]; because it was violated for the ratio and rebound hyperglycemia, HRs were also estimated for 0–6, 6–12, 12–24 and 6–24 h among episodes at risk at the start of each interval.

Missing covariate values, mainly HbA1c (6.5%), were imputed 20 times by chained equations that included all covariates, the exposure, the outcome indicators and the Nelson–Aalen cumulative hazards of the primary outcomes [30]; estimates were combined with Rubin’s rules [31]. The hypoglycemia model had about five events per estimated parameter, within the range supported by simulation studies [32]; a parsimonious model with about ten events per parameter was also fitted, and clinically significant hypoglycemia (55 events) was analyzed with a minimal adjustment set (Additional file 1: Supplementary Methods).

In secondary analyses, we estimated the associations of glargine administration at least 2 h before discontinuation, reflecting the American Diabetes Association recommendation of administration 2 h before [4], and of an oral diet at discontinuation with the primary outcomes; for timing, models were adjusted only for characteristics preceding the dose, timing was also analyzed in five categories, and the comparison of doses given at least 2 h with those given 0 to <2 h before discontinuation excluded doses given after discontinuation. Effect modification of the association between the ratio and each primary outcome was assessed with interaction terms for diabetes, HbA1c of 8.0% or higher (among episodes with a measured value), insulin use before admission (among episodes with known status), cardiac surgery, an oral diet, glargine administration at least 2 h before discontinuation, corticosteroid therapy, age of 65 years or older, and sex; the interaction with oral diet was also examined for 0–6 and 6–24 h. To express the dose–response relationship in absolute terms, risks were standardized to the cohort at selected ratios with the parametric g-formula; because hazards were not proportional for rebound hyperglycemia, its risk was modeled separately for 0–6 and 6–24 h, and 95% CIs were obtained by patient-level bootstrap resampling. The shape of the relationship was also examined with three and five spline knots, with six ratio categories and, for rebound hyperglycemia, among episodes at risk 6 h after discontinuation.

Twenty-one sensitivity analyses (Additional file 1: Supplementary Methods) addressed missing data; informative censoring by ICU discharge or resumption of IV insulin, including inverse probability of censoring weighting with pooled logistic models, reported as odds ratios [33], and restriction to episodes with at least 24 h of potential ICU follow-up; estimation of the IV requirement over up to 24 h or over the time the infusion was running; restrictions of the study population and of the exposure distribution; calendar period; model specification; and the definition of hypoglycemia. The glargine dose per kilogram of body weight was examined as an alternative exposure, and the E-value quantified the strength of unmeasured confounding needed to explain away the hypoglycemia association [34]. Tests were two-sided, with P < 0.05 considered statistically significant; secondary, subgroup and sensitivity analyses were considered exploratory and were not adjusted for multiplicity. HRs are shown to three decimal places when a confidence limit rounds to 1.00. Analyses were performed in Python 3.12.9 (NumPy, pandas, SciPy and scikit-learn); Cox models were fitted with custom code, and the fully adjusted models were reproduced with the lifelines package (Additional file 1: Supplementary Methods).

## Results

### Characteristics of transition episodes

Of 13,824 IV insulin infusion episodes in 11,141 patients, 6,220 transition episodes in 5,924 patients met the eligibility criteria (Fig. 1); 237 patients contributed more than one episode. Most episodes followed cardiac surgery (87.5%) and took place in the cardiac vascular ICU (92.5%). The median age was 67 years (IQR 60–74), 70.9% of episodes involved men, and 46.0% involved patients with diabetes (Table 1; Additional file 1: Table S4). The first glargine dose was a median of 20 U (IQR 10–30), or 0.24 U/kg (0.16–0.34), given a median of 1.8 h (IQR 1.1–2.0) before discontinuation; 30.4% of doses were given at least 2 h before discontinuation. The median glargine-to-IV dose ratio was 0.41 (IQR 0.30–0.46), and 1,048 episodes (16.8%) had a ratio of 0.417, corresponding to a dose of 10 times the mean hourly infusion rate (Additional file 1: Table S5). Compared with the reference category, episodes with a ratio of 0.60 or higher received a larger glargine dose (median 30 vs 20 U; 0.32 vs 0.25 U/kg) against a lower and more steeply falling estimated IV total daily dose (32.6 vs 48.0 U; rate trend 0.54 vs 0.82), and more often involved diabetes (57.8% vs 38.0%), insulin use before admission (27.9% vs 11.6%) and an oral diet at discontinuation (20.8% vs 9.6%).

**Table 1.**
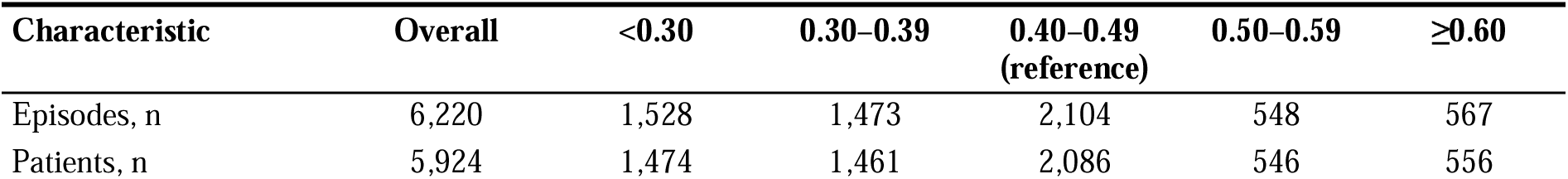
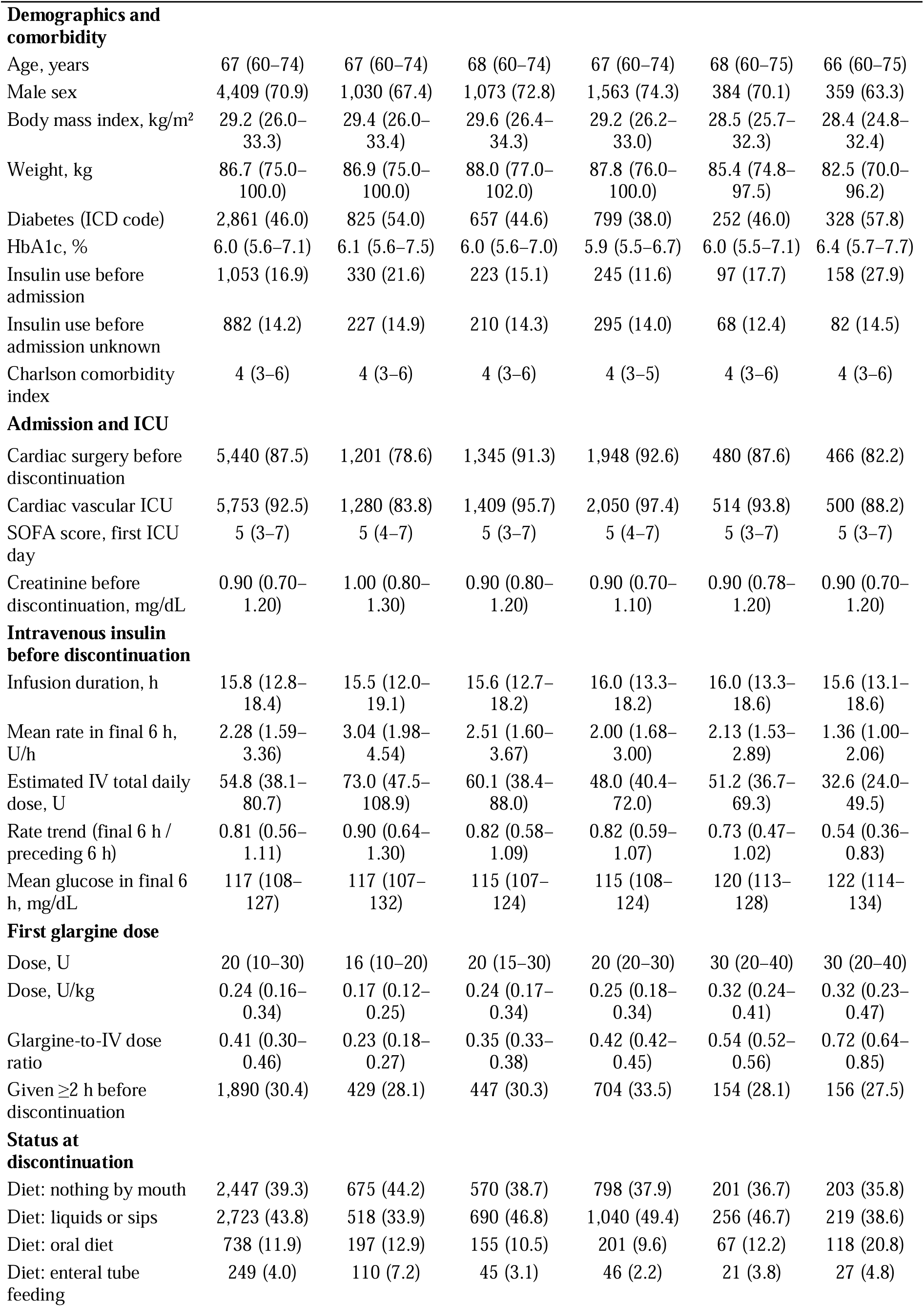
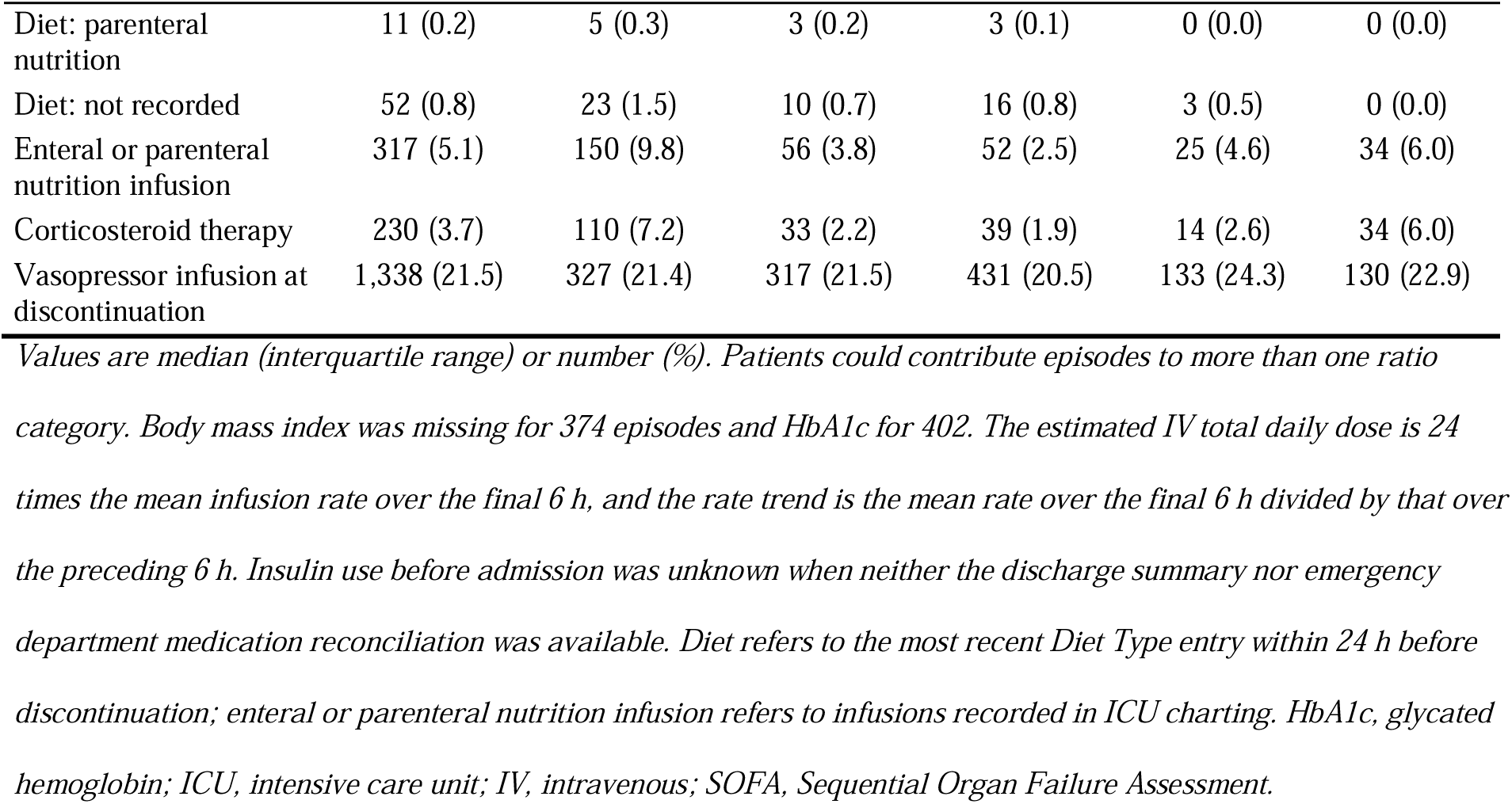
Characteristics of transition episodes by glargine-to-intravenous dose ratio.

### Glargine-to-IV dose ratio and glycemic outcomes

During ICU follow-up (median potential follow-up 15.2 h [IQR 8.9–24.0] within the 24-h window; complete 24-h follow-up in 44.1% of episodes), rebound hyperglycemia occurred in 2,109 episodes (Kaplan–Meier cumulative incidence 44.4%, 95% CI 42.8–45.9) and hypoglycemia in 211 (cumulative incidence 6.4%, 5.5–7.4), a median of 5.3 h and 10.8 h after discontinuation, respectively (Table 2; Additional file 1: Table S6). The unadjusted cumulative incidence of rebound hyperglycemia was lowest in the reference category (35.4%) and higher at both lower and higher ratios, whereas that of hypoglycemia increased from 5.8% in the reference category to 10.6% at ratios of 0.60 or higher. After adjustment, each 0.1 increase in the ratio was associated with a 4% lower hazard of rebound hyperglycemia (HR 0.96, 95% CI 0.93–0.99) and an 11% higher hazard of hypoglycemia (HR 1.11, 1.04–1.18); the U-shaped unadjusted pattern for rebound hyperglycemia was largely explained by differences in patient characteristics (estimates for all covariates in Additional file 1: Table S7). Ratios of 0.60 or higher were associated with more hypoglycemia than the reference category (HR 1.76, 1.09–2.83), with an HR of 0.85 (0.70–1.02) for rebound hyperglycemia. Higher ratios were also associated with less severe hyperglycemia (HR per 0.1 increase 0.93, 0.89–0.98) and less frequent resumption of IV insulin (HR 0.90, 0.83–0.98); clinically significant hypoglycemia (55 events) showed a consistent direction (minimally adjusted HR per 0.1 increase 1.08, 0.99–1.19; Additional file 1: Table S8).

**Table 2.**
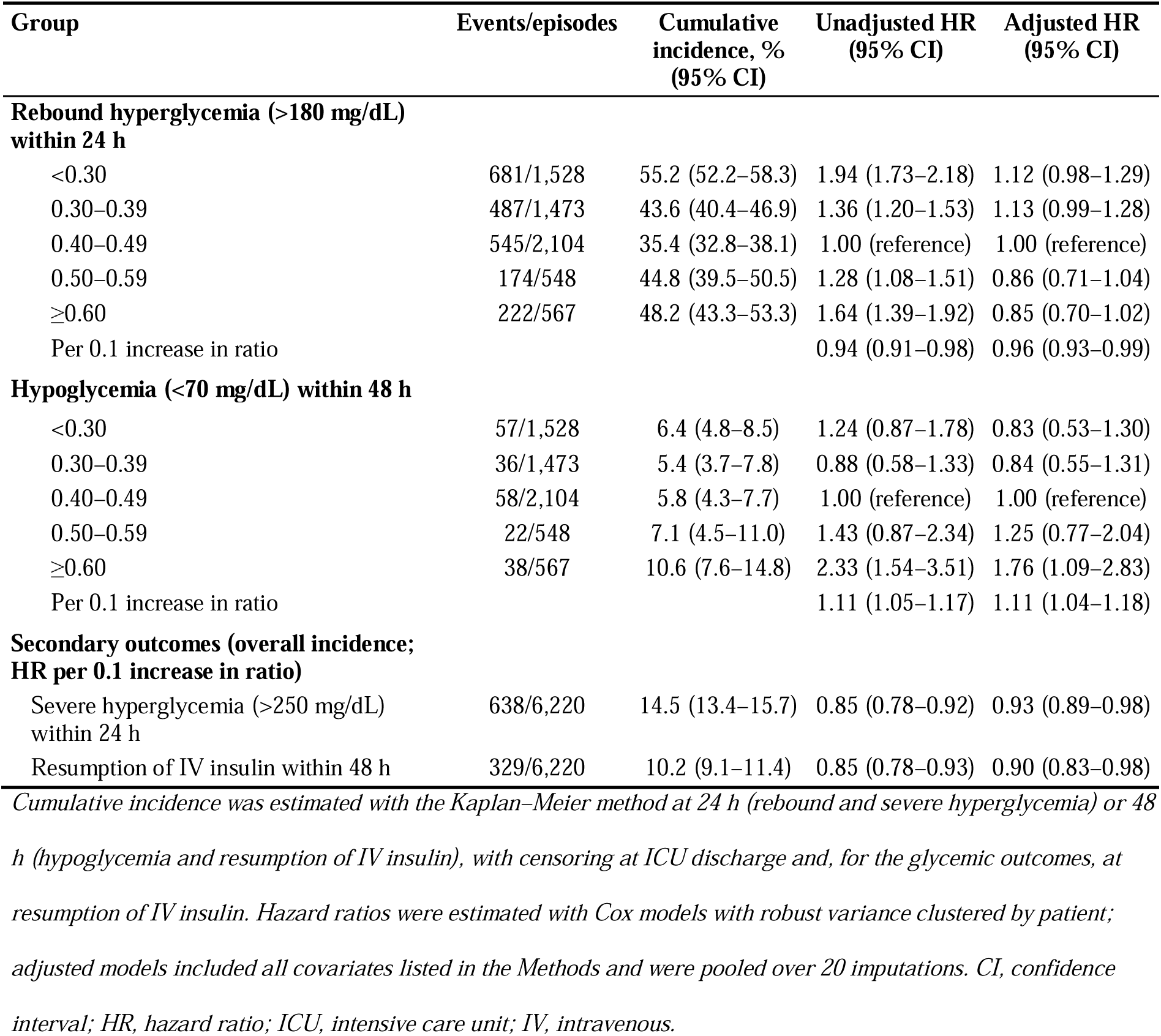
Glycemic outcomes by glargine-to-intravenous dose ratio.

| Group | Events/episodes | Cumulative incidence, % (95% CI) | Unadjusted HR (95% CI) | Adjusted HR (95% CI) |
| --- | --- | --- | --- | --- |
| **Rebound hyperglycemia (>180 mg/dL) within 24 h** |  |  |  |  |
| <0.30 | 681/1,528 | 55.2 (52.2–58.3) | 1.94 (1.73–2.18) | 1.12 (0.98–1.29) |
| 0.30–0.39 | 487/1,473 | 43.6 (40.4–46.9) | 1.36 (1.20–1.53) | 1.13 (0.99–1.28) |
| 0.40–0.49 | 545/2,104 | 35.4 (32.8–38.1) | 1.00 (reference) | 1.00 (reference) |
| 0.50–0.59 | 174/548 | 44.8 (39.5–50.5) | 1.28 (1.08–1.51) | 0.86 (0.71–1.04) |
| ≥0.60 | 222/567 | 48.2 (43.3–53.3) | 1.64 (1.39–1.92) | 0.85 (0.70–1.02) |
| Per 0.1 increase in ratio |  |  | 0.94 (0.91–0.98) | 0.96 (0.93–0.99) |
| **Hypoglycemia (<70 mg/dL) within 48 h** |  |  |  |  |
| <0.30 | 57/1,528 | 6.4 (4.8–8.5) | 1.24 (0.87–1.78) | 0.83 (0.53–1.30) |
| 0.30–0.39 | 36/1,473 | 5.4 (3.7–7.8) | 0.88 (0.58–1.33) | 0.84 (0.55–1.31) |
| 0.40–0.49 | 58/2,104 | 5.8 (4.3–7.7) | 1.00 (reference) | 1.00 (reference) |
| 0.50–0.59 | 22/548 | 7.1 (4.5–11.0) | 1.43 (0.87–2.34) | 1.25 (0.77–2.04) |
| ≥0.60 | 38/567 | 10.6 (7.6–14.8) | 2.33 (1.54–3.51) | 1.76 (1.09–2.83) |
| Per 0.1 increase in ratio |  |  | 1.11 (1.05–1.17) | 1.11 (1.04–1.18) |
| **Secondary outcomes (overall incidence; HR per 0.1 increase in ratio)** |  |  |  |  |
| Severe hyperglycemia (>250 mg/dL) within 24 h | 638/6,220 | 14.5 (13.4–15.7) | 0.85 (0.78–0.92) | 0.93 (0.89–0.98) |
| Resumption of IV insulin within 48 h | 329/6,220 | 10.2 (9.1–11.4) | 0.85 (0.78–0.93) | 0.90 (0.83–0.98) |

Cumulative incidence was estimated with the Kaplan–Meier method at 24 h (rebound and severe hyperglycemia) or 48 h (hypoglycemia and resumption of IV insulin), with censoring at ICU discharge and, for the glycemic outcomes, at resumption of IV insulin. Hazard ratios were estimated with Cox models with robust variance clustered by patient; adjusted models included all covariates listed in the Methods and were pooled over 20 imputations. CI, confidence interval; HR, hazard ratio; ICU, intensive care unit; IV, intravenous.

### Dose–response relationship and absolute risks

In spline analysis, the hazard of rebound hyperglycemia decreased as the ratio increased (P = 0.008 for the overall association; P = 0.12 for nonlinearity), with HRs relative to a ratio of 0.40 of 1.11 (1.04–1.19) at 0.30, 0.87 (0.79–0.96) at 0.60 and 0.85 (0.75–0.97) at 0.80 (Fig. 2). The hazard of hypoglycemia increased with the ratio (P = 0.016; P = 0.47 for nonlinearity), with HRs of 0.81 (0.66–0.99) at 0.30, 1.318 (1.004–1.729) at 0.60 and 1.52 (1.09–2.13) at 0.80. Standardized to the cohort, the estimated risks of both outcomes changed gradually across the range of ratios (Table 3). Relative to a ratio of 0.40, a ratio of 0.60 corresponded to 2.7 (0.6–4.9) fewer episodes of rebound hyperglycemia and 1.7 (−0.2 to 3.5) more episodes of hypoglycemia per 100 transitions, and a ratio of 0.30 to 2.4 (0.8–4.0) more and 1.1 (0.1–2.0) fewer; each 0.1 increase between 0.30 and 0.80 corresponded to 1.4 to 2.3 fewer episodes of rebound hyperglycemia for each additional episode of hypoglycemia. Among episodes at risk 6 h after discontinuation, the hazard of rebound hyperglycemia tended to decrease further above a ratio of 0.60 (HR 0.89, 0.76–1.04, for 0.80 vs 0.60), and the shape of both associations was similar with three or five spline knots and with six ratio categories (Additional file 1: Table S9).

**Fig. 2.**
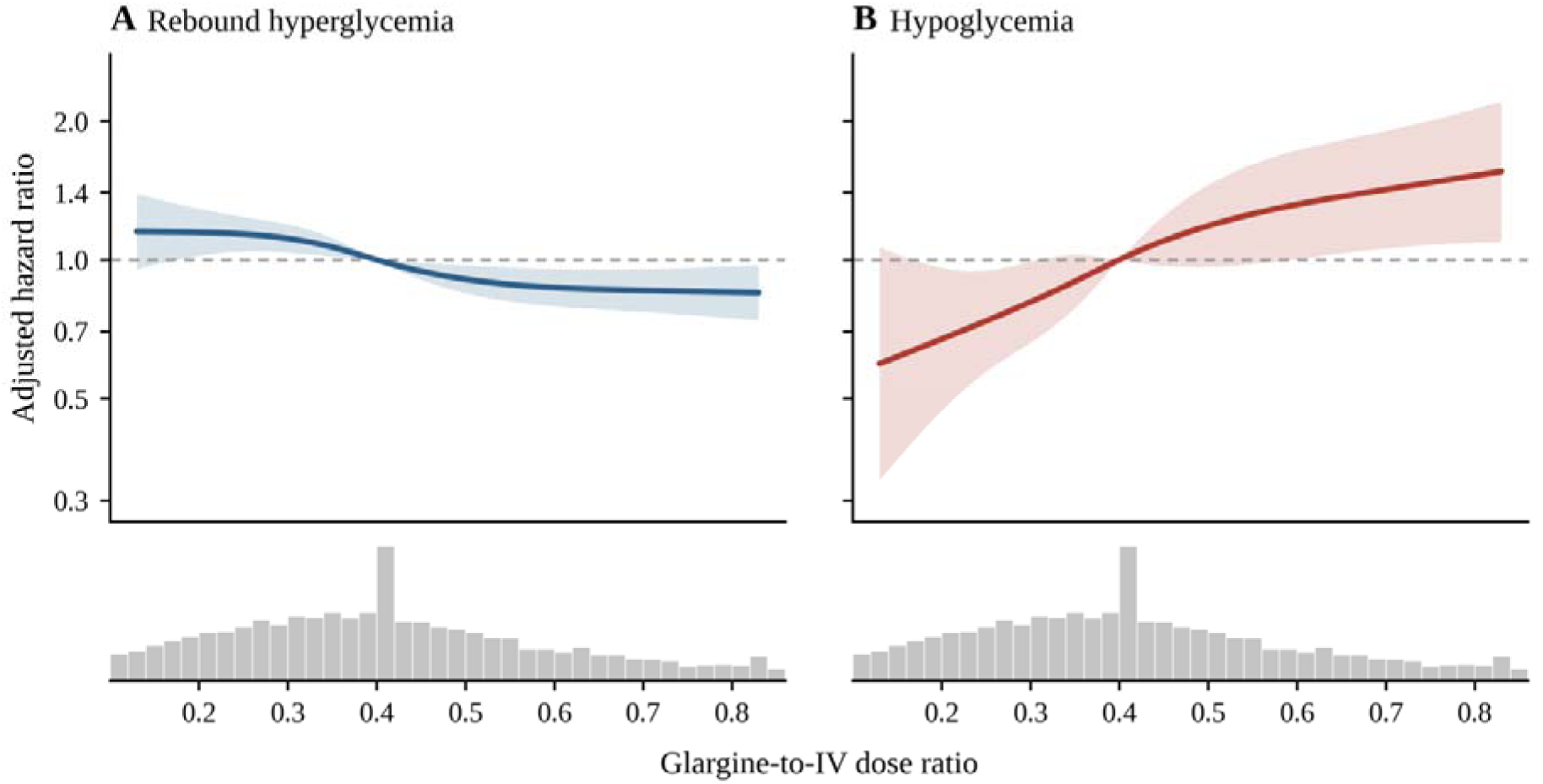
Association between the glargine-to-intravenous dose ratio and the primary outcomes. Adjusted hazard ratios (solid lines) and 95% confidence intervals (shaded bands) for (A) rebound hyperglycemia (glucose >180 mg/dL within 24 h) and (B) hypoglycemia (glucose <70 mg/dL within 48 h), relative to a ratio of 0.40, on a logarithmic scale shared by both panels. For rebound hyperglycemia, whose association with the ratio emerged after the first 6 h, the curve represents an average over the 24-h window. The ratio was modeled with restricted cubic splines with knots at 0.16, 0.35, 0.42 and 0.70; models were adjusted for all covariates listed in the Methods and pooled over 20 imputations. The dashed horizontal line indicates a hazard ratio of 1. The histograms below the panels show the distribution of the ratio on a square-root scale (bin width 0.02); 233 episodes with ratios below 0.10 or of 0.86 or higher are not shown. Curves are displayed between the 2.5th and 97.5th percentiles of the ratio (0.12–0.83). P values for the overall association and for nonlinearity: (A) P = 0.008 and P = 0.12; (B) P = 0.016 and P = 0.47.

**Table 3.**
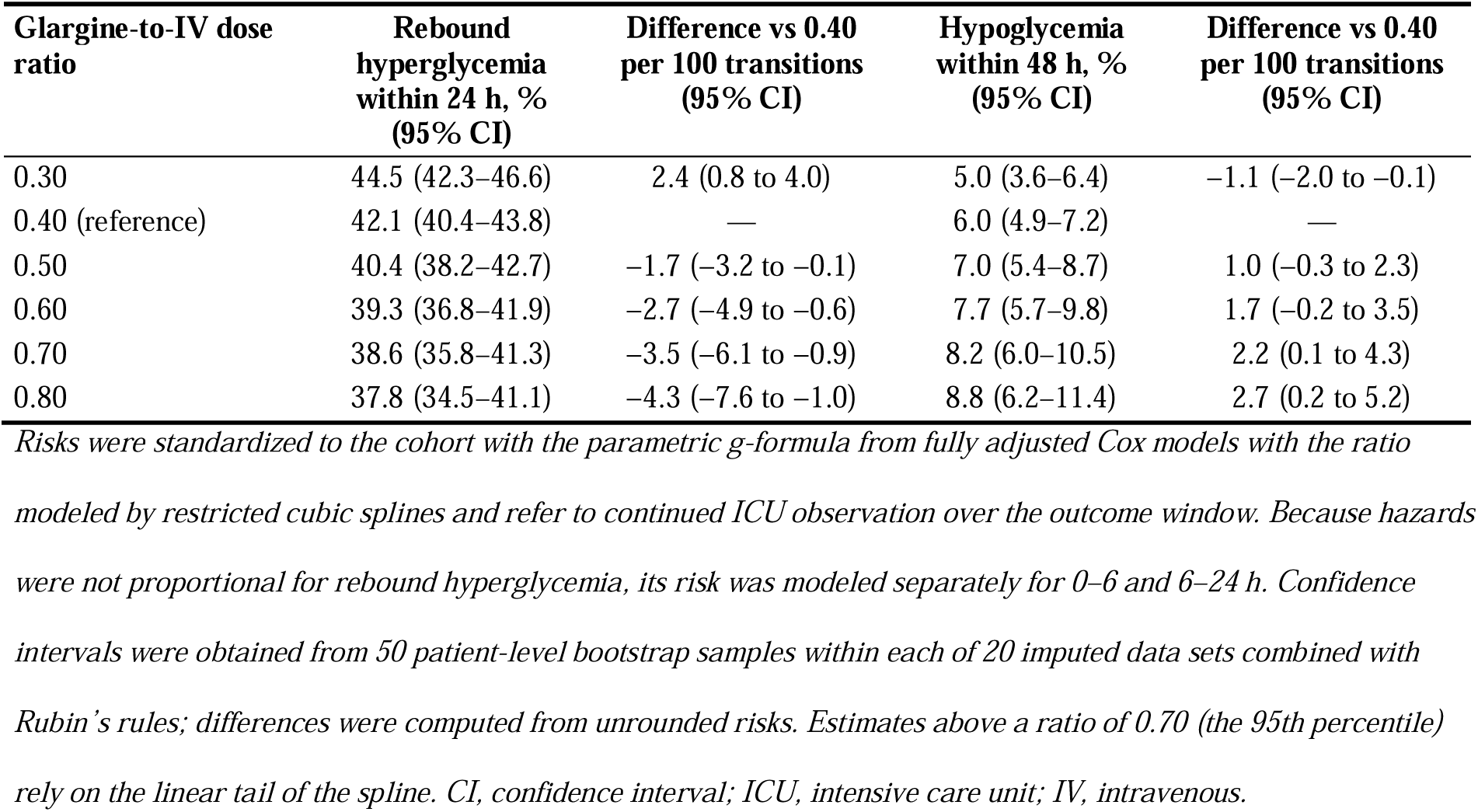
Standardized risks of the primary outcomes at selected glargine-to-intravenous dose ratios.

| **Glargine-to-IV dose ratio** | **Rebound hyperglycemia within 24 h, % (95% CI)** | **Difference vs 0.40 per 100 transitions (95% CI)** | **Hypoglycemia within 48 h, % (95% CI)** | **Difference vs 0.40 per 100 transitions (95% CI)** |
| --- | --- | --- | --- | --- |
| 0.30 | 44.5 (42.3–46.6) | 2.4 (0.8 to 4.0) | 5.0 (3.6–6.4) | –1.1 (–2.0 to –0.1) |
| 0.40 (reference) | 42.1 (40.4–43.8) | — | 6.0 (4.9–7.2) | — |
| 0.50 | 40.4 (38.2–42.7) | –1.7 (–3.2 to –0.1) | 7.0 (5.4–8.7) | 1.0 (–0.3 to 2.3) |
| 0.60 | 39.3 (36.8–41.9) | –2.7 (–4.9 to –0.6) | 7.7 (5.7–9.8) | 1.7 (–0.2 to 3.5) |
| 0.70 | 38.6 (35.8–41.3) | –3.5 (–6.1 to –0.9) | 8.2 (6.0–10.5) | 2.2 (0.1 to 4.3) |
| 0.80 | 37.8 (34.5–41.1) | –4.3 (–7.6 to –1.0) | 8.8 (6.2–11.4) | 2.7 (0.2 to 5.2) |

*Risks were standardized to the cohort with the parametric g-formula from fully adjusted Cox models with the ratio modeled by restricted cubic splines and refer to continued ICU observation over the outcome window. Because hazards were not proportional for rebound hyperglycemia, its risk was modeled separately for 0–6 and 6–24 h. Confidence intervals were obtained from 50 patient-level bootstrap samples within each of 20 imputed data sets combined with Rubin's rules; differences were computed from unrounded risks. Estimates above a ratio of 0.70 (the 95th percentile) rely on the linear tail of the spline. CI, confidence interval; ICU, intensive care unit; IV, intravenous.*

### Time course of rebound hyperglycemia

The association between the ratio and rebound hyperglycemia was not constant over time (P = 0.020 for nonproportionality; Additional file 1: Table S10). Rebound in the first 6 h after discontinuation, which accounted for 57.6% of rebound events, was not associated with the ratio overall (HR per 0.1 increase 0.99, 0.95–1.02; ratios of 0.60 or higher vs 0.40–0.49, 0.94, 0.74–1.19), whereas rebound between 6 and 24 h became less frequent as the ratio increased (HR per 0.1 increase 0.91, 0.86–0.96; 12–24 h: 0.87, 0.81–0.94). An oral diet was associated with more rebound hyperglycemia in both periods (HR 1.53, 1.28–1.83, within 6 h and 1.42, 1.15–1.75, between 6 and 24 h). The proportional hazards assumption was not rejected for hypoglycemia (P = 0.52).

### Timing of glargine and nutritional status

In models adjusted for characteristics preceding the dose, glargine given at least 2 h before discontinuation, compared with 0 to <2 h before, was associated with less rebound hyperglycemia (HR 0.86, 0.77–0.96) without a significant difference in hypoglycemia (HR 1.14, 0.85–1.53) (Additional file 1: Table S11). Relative to doses given 1 to <2 h before discontinuation, the HR for rebound hyperglycemia within 24 h was 1.05 (0.87–1.28) for doses given after discontinuation, 0.97 (0.85–1.11) for 0 to <1 h, 0.87 (0.77–0.98) for 2 to <3 h and 0.82 (0.67–1.01) for 3 h or more before discontinuation, whereas the pattern within the first 6 h was irregular; doses given 3 h or more before discontinuation had a higher but imprecise HR for hypoglycemia (1.53, 0.90–2.61). An oral diet at discontinuation was associated with more rebound hyperglycemia (HR 1.49, 1.30–1.71) but not with hypoglycemia (HR 0.89, 0.56–1.43). The unadjusted cumulative incidence of rebound hyperglycemia was 64.8% with an oral diet and 88.1% with enteral tube feeding, which also had the highest unadjusted incidence of hypoglycemia (11.0%) (Additional file 1: Table S12).

### Subgroup and sensitivity analyses

The association between the ratio and hypoglycemia was consistent across predefined subgroups (all P for interaction > 0.05), with HRs per 0.1 increase ranging from 1.04 to 1.22 (Fig. 3; Additional file 1: Table S13). For rebound hyperglycemia, the association differed by nutritional status (P for interaction = 0.011), and this difference was confined to the first 6 h: within 6 h, the HR per 0.1 increase was 0.956 (0.914–1.000) without and 1.04 (0.99–1.09) with an oral diet (P for interaction = 0.003), whereas between 6 and 24 h it was 0.91 (0.86–0.97) and 0.90 (0.83–0.98) (P for interaction = 0.88). No other interaction was statistically significant.

**Fig. 3.**
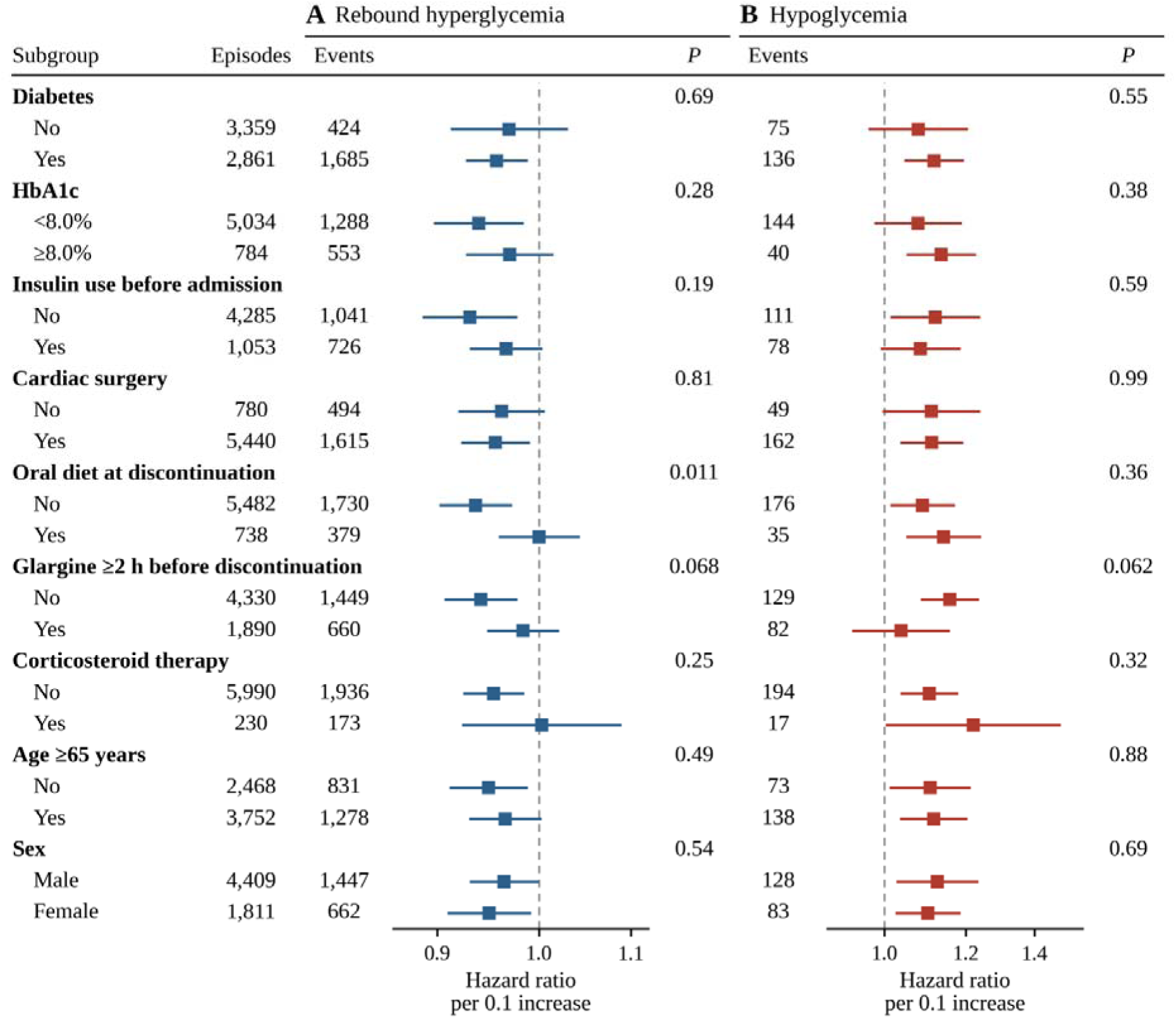
Association between the glargine-to-intravenous dose ratio and the primary outcomes in predefined subgroups. Adjusted hazard ratios and 95% confidence intervals per 0.1 increase in the ratio for (A) rebound hyperglycemia and (B) hypoglycemia, on a logarithmic scale, estimated from Cox models including an interaction between the ratio and each subgrouping variable, adjusted for all covariates listed in the Methods and pooled over 20 imputations. The HbA1c subgroup analysis was restricted to episodes with a measured HbA1c (n = 5,818), and the analysis by insulin use before admission to episodes with known status (n = 5,338). Numbers of episodes and events and P values for interaction are shown for each subgroup. HbA1c, glycated hemoglobin.

Both associations were consistent in direction across the 21 sensitivity analyses (Additional file 1: Table S14). HRs per 0.1 increase ranged from 1.09 to 1.21 for hypoglycemia and from 0.93 to 0.96 for rebound hyperglycemia, except for attenuation of the rebound association with linear covariate terms (HR 0.98, 0.95–1.01), a specification that fitted the data less well; odds ratios after inverse probability of censoring weighting were 1.13 (1.06–1.20) and 0.96 (0.93–0.99). Estimates for ratios of 0.60 or higher were less precise (hypoglycemia HRs 1.39–2.42). Interruptions of 15 min or longer in the final 6 h were more common at higher ratios (57.1% of episodes at ratios of 0.60 or higher vs 8.1% in the reference category); recalculating the ratio over the time the infusion was running reduced the number of episodes with ratios of 0.60 or higher from 567 to 286, and the HRs per 0.1 increase were 1.10 (1.03–1.18) for hypoglycemia and 0.964 (0.934–0.996) for rebound hyperglycemia. The hypoglycemia association persisted when follow-up was limited to 24 h (HR 1.12, 1.05–1.20) and when events preceded by short-acting insulin within 4 h were censored (HR 1.09, 1.02–1.17); such insulin preceded hypoglycemia in 9 of 38 episodes (23.7%) at ratios of 0.60 or higher, 4 of 58 (6.9%) in the reference category and 14.0–19.4% in the other categories.

The glargine dose per kilogram showed associations in the same direction (per 0.1 U/kg: HR 0.93 [0.90–0.98] for rebound hyperglycemia and 1.17 [1.04–1.30] for hypoglycemia), and the E-value for each 0.1 increase in the ratio was 1.46 (1.26 for the lower confidence limit). During the first 24 h, glucose was measured a median of 0.25–0.26 times per hour in each ratio category, short-acting insulin was given in 79.9% of episodes, and the mean proportion of glucose values within 70–180 mg/dL was highest in the reference category (86.6%); additional basal insulin within 48 h was given in 30.3% of episodes at ratios below 0.30 or of 0.60 or higher and in 17.9% in the reference category (Additional file 1: Table S15).

## Discussion

In this cohort of 6,220 transitions from IV insulin to subcutaneous glargine, most of them after cardiac surgery, higher glargine doses relative to the preceding IV requirement were associated with less rebound hyperglycemia and more hypoglycemia. The association with rebound hyperglycemia emerged after the first 6 h and was accompanied by less severe hyperglycemia and less frequent resumption of IV insulin, suggesting that it was not confined to single glucose values just above 180 mg/dL. In absolute terms, the two outcomes changed gradually and in opposite directions across the range of ratios, with 1.4 to 2.3 fewer episodes of rebound hyperglycemia for each additional episode of hypoglycemia per 0.1 increase and no clear threshold.

For hospitalized patients, the American Diabetes Association suggests a subcutaneous total daily dose of about 60% of that estimated from the infusion rate, half of it as basal insulin, which implies a basal fraction of about 0.30 [4]. Most transitions in our cohort exceeded this fraction (median 0.41), possibly because 83.1% of transitions occurred when patients were receiving nothing by mouth or only liquids, when the nutritional half of such a regimen would not be given; relative to 0.40, a ratio of 0.30 corresponded to 2.4 more episodes of rebound hyperglycemia and 1.1 fewer episodes of hypoglycemia per 100 transitions. The randomized trials of conversion doses were small and not designed to quantify this trade-off, and none identified a clearly superior basal fraction [9–11]; in one, hypoglycemia occurred only with the two higher doses of detemir [11], consistent with our findings. A retrospective ICU study that used the same estimate of the IV requirement found the highest proportion of glucose values in range with initial basal doses of 50–59% [12]; in our cohort, a ratio of 0.50 rather than 0.40 corresponded to 1.7 fewer episodes of rebound hyperglycemia and 1.0 more episode of hypoglycemia per 100 transitions (Table 3), a trade-off that the proportion of values in range may not capture. High ratios reflected both larger glargine doses and a low, falling IV requirement, often with interruptions of the infusion in the final hours; the median dose was 30 U in both the 0.50–0.59 and the higher category, suggesting that the dose was often not reduced when the requirement fell, and the glargine dose per kilogram was associated with both outcomes in the same direction as the ratio.

Glargine has a slow onset and a flat, prolonged action profile [35], so glucose control in the first hours after the infusion is stopped may depend more on the overlap between the two insulins than on the glargine dose. Accordingly, early rebound hyperglycemia was not associated with the ratio overall, whereas later rebound was less frequent at higher ratios. Administration of glargine at least 2 h before discontinuation, in line with the American Diabetes Association recommendation to give basal insulin 2 h before the infusion is stopped [4], was associated with less rebound hyperglycemia, consistent with the finding that basal insulin started during the infusion prevents rebound hyperglycemia [8]; this association may partly reflect planned, protocol-driven transitions, and the imprecisely higher HR for hypoglycemia with doses given 3 h or more before discontinuation suggests that timing should be evaluated together with dose. Hypoglycemia occurred later, when glargine action was established and insulin requirements may fall as stress hyperglycemia resolves [1]; its association with the ratio was robust to the estimation of the IV requirement, the handling of infusion interruptions and the length of follow-up, and short-acting insulin given before hypoglycemia did not fully explain it.

Nutritional status was strongly associated with rebound hyperglycemia. Patients eating an oral diet had a 49% higher hazard of rebound hyperglycemia, consistent with prandial hyperglycemia that basal insulin is not intended to cover, and in line with the superiority of basal–bolus regimens over sliding-scale insulin [36] and with guidance for non-critical care settings favoring scheduled over correctional-only insulin [37]. The association with the ratio differed by diet only in the first 6 h (borderline without and absent with an oral diet), not thereafter. Patients receiving enteral tube feeding had the highest unadjusted incidence of both rebound hyperglycemia and hypoglycemia, suggesting that insulin matched to, and adjustable with, the feeding deserves evaluation in these patients.

Strengths of this study include the number of transitions and hypoglycemia events, reconciliation of two medication records with exact dose times, flexible adjustment for the infusion rate and its trend, which are mathematically coupled to the exposure, absolute risk estimates that allow for the changing association over time, and multiple sensitivity analyses.

This study has limitations. First, residual confounding by indication is possible, because doses and their timing may have been chosen on the basis of factors not captured in the data, such as anticipated oral intake or a planned transition; an unmeasured confounder would need to be associated with both the ratio and hypoglycemia by a risk ratio of 1.46 per 0.1 increase in the ratio (1.26 for the confidence limit) to explain away the association. Second, the data came from a single center, where 87.5% of transitions followed cardiac surgery and about half of the reference category received a dose of 10 times the mean hourly infusion rate, so the findings and the distribution of ratios may differ in medical ICUs and at other centers. Third, follow-up was limited to the ICU, and hypoglycemia after ICU discharge may have been missed, although inverse probability of censoring weighting and restriction to episodes with at least 24 h of potential follow-up gave similar results. Fourth, the exposure was derived from charted infusion records, in which interruptions counted as zero rate and the final 6 h included a median of 1.8 h after the glargine dose, when its effect on the infusion rate is likely small; the association per 0.1 increase was robust to the handling of interruptions and to a 24-h estimate of the requirement, and intermittent glucose measurement, whose frequency was similar across ratio categories, may have led to largely non-differential underascertainment of both outcomes.

In patients similar to those studied, the initial glargine dose involves a gradual trade-off rather than a threshold: the basal fraction of about 0.30 implied by current guidance favors less hypoglycemia, higher fractions favor less rebound hyperglycemia, and the preferred fraction depends on the relative weight given to the two outcomes. Reducing the dose when the IV requirement is falling, giving glargine before the infusion is stopped and matching insulin to nutrition are hypotheses for a randomized trial comparing basal fractions of about 0.30 and 0.50, using continuous glucose monitoring that extends beyond ICU discharge.

## Conclusions

In critically ill adults transitioned from IV insulin to glargine, mostly after cardiac surgery, higher glargine doses relative to the preceding IV requirement were associated with less rebound hyperglycemia after the first 6 h and with more hypoglycemia, a trade-off that changed gradually across the range used in practice. Which basal fraction is preferable depends on the relative weight given to these outcomes; a randomized trial that includes the fraction of about 0.30 implied by current guidance is warranted.

## Data Availability

The datasets analyzed during the current study are available in the PhysioNet repository: MIMIC-IV version 3.1, https://doi.org/10.13026/kpb9-mt58; MIMIC-IV-Note version 2.2, https://doi.org/10.13026/1n74-ne17; and MIMIC-IV-ED version 2.2, https://doi.org/10.13026/5ntk-km72. Access is restricted to credentialed users who complete the required training and sign the data use agreement, and the authors are not permitted to share the data. The analytic code and the exact cohort-extraction queries used in this study are openly available in the Zenodo repository (https://doi.org/10.5281/zenodo.23004506).

## List of abbreviations

CI: Confidence interval
DKA: Diabetic ketoacidosis
eMAR: Electronic medication administration record
HbA1c: Glycated hemoglobin
HHS: Hyperosmolar hyperglycemic state
HR: Hazard ratio
ICU: Intensive care unit
IQR: Interquartile range
IV: Intravenous
MIMIC-IV: Medical Information Mart for Intensive Care IV
RECORD: REporting of studies Conducted using Observational Routinely-collected health Data
SOFA: Sequential Organ Failure Assessment
SQL: Structured Query Language
STROBE: Strengthening the Reporting of Observational Studies in Epidemiology

## Declarations

## Ethics approval and consent to participate

This study is a secondary analysis of a de-identified, publicly available critical care database, MIMIC-IV, including its linked emergency department (MIMIC-IV-ED) and clinical note (MIMIC-IV-Note) modules, accessed through PhysioNet by a credentialed investigator who completed the required human-subjects research training and signed the data use agreement. The collection of patient information and the creation of the database were reviewed by the Institutional Review Board at Beth Israel Deaconess Medical Center, which approved the data sharing initiative and waived the requirement for individual informed consent because the data are de-identified. No additional ethics approval was required for this secondary analysis. All methods were performed in accordance with the Declaration of Helsinki and relevant guidelines and regulations.

## Consent for publication

Not applicable.

## Clinical trial number

Not applicable.

## Availability of data and materials

The datasets analyzed during the current study are available in the PhysioNet repository: MIMIC-IV version 3.1, https://doi.org/10.13026/kpb9-mt58 [15]; MIMIC-IV-Note version 2.2, https://doi.org/10.13026/1n74-ne17 [17]; and MIMIC-IV-ED version 2.2, https://doi.org/10.13026/5ntk-km72 [18]. Access is restricted to credentialed users who complete the required training and sign the data use agreement, and the authors are not permitted to share the data. The analytic code and the exact cohort-extraction queries used in this study are openly available in the Zenodo repository (https://doi.org/10.5281/zenodo.23004506).

## Competing interests

The authors declare that they have no competing interests.

## Funding

This research received no specific grant from any funding agency in the public, commercial, or not-for-profit sectors.

## Authors’ contributions

LS conceived and designed the study, extracted and curated the data, performed the statistical analyses, interpreted the results and drafted the manuscript. YL contributed to data curation, verified the data extraction and analyses and critically revised the manuscript. LW contributed to the literature review and the clinical interpretation of the results and critically revised the manuscript. WS conceived the study, supervised the work, contributed to the interpretation of the results and critically revised the manuscript. All authors read and approved the final manuscript.

## Acknowledgements

Not applicable.

## Additional files

### Additional file 1

*File format:* Microsoft Word document (.docx)

*Title of data:* Supplementary Methods and Tables S1–S15

*Description of data:* Detailed definitions and analytical methods; data elements and codes; reconciliation of glargine records; covariate definitions and missing data; complete baseline characteristics; distribution of the glargine-to-intravenous dose ratio; cumulative incidence of the primary outcomes over time; estimates for all covariates; results for the secondary outcomes; shape of the dose–response relationship; assessment of proportional hazards and interval-specific estimates; timing of glargine; outcomes and insulin use by nutritional status; subgroup analyses; sensitivity analyses; and glycemic control and insulin use after discontinuation.

### Additional file 2

*File format:* Microsoft Word document (.docx)

*Title of data:* RECORD checklist

*Description of data:* Completed RECORD checklist, which incorporates the STROBE items, indicating where each item is reported in the manuscript.

## Notes

### Competing Interest Statement

The authors have declared no competing interest.

### Author Declarations

The Institutional Review Board of the Beth Israel Deaconess Medical Center waived ethical approval for this work. The collection of patient information and creation of the MIMIC-IV database were reviewed by the Institutional Review Board of the Beth Israel Deaconess Medical Center, which approved the data sharing initiative and waived the requirement for individual informed consent because the data are de-identified. This study was a secondary analysis of de-identified MIMIC-IV data accessed through PhysioNet under the required data use agreement, and no additional ethics approval was required for this secondary analysis.

