## Supplementary Methods and Supplementary Tables S1-S15 for "Glargine-to-intravenous insulin dose ratio, rebound hyperglycemia and hypoglycemia after discontinuation of insulin infusion in critically ill adults: a retrospective cohort study"

**Additional file 1**

Contents: Supplementary Methods; Tables S1–S15. Abbreviations are as defined in the main manuscript. Reference numbers refer to the reference list of the main manuscript, except reference S1, which is listed at the end of the Supplementary Methods.

### ***Supplementary Methods***

**Data and extraction.** Data were extracted from MIMIC-IV version 3.1 (hosp and icu modules), MIMIC-IV-ED version 2.2 and MIMIC-IV-Note version 2.2 [14–18] with Structured Query Language (SQL) on Google BigQuery; first-day Sequential Organ Failure Assessment (SOFA) scores and Charlson comorbidity indices were taken from the derived tables of the MIMIC Code Repository [22–25]. Queries returned one row per infusion episode (identified by ICU stay and time of discontinuation), ICU stay or admission, and all 13,824 infusion episodes were matched across files. Data elements and codes are listed in Table S1. The SQL queries and analysis code are available in the Zenodo repository cited in the main manuscript.

**Infusion episodes and time zero.** Charted records of IV regular insulin with a positive rate, excluding records flagged as rewritten, were ordered by start time within each ICU stay, and a new episode began when a record started more than 6 h after the latest end time of all preceding records. The latest end time defined discontinuation (time zero), and episodes shorter than 6 h were not analyzed. The start of the next episode in the same ICU stay defined resumption of IV insulin, which could therefore occur only more than 6 h after discontinuation. Basal insulin comprised glargine, neutral protamine Hagedorn (NPH) insulin, premixed insulin and detemir; exclusions are shown in Fig. 1 of the main text.

**Exposure.** The first basal insulin administration from 12 h before to 1 h after discontinuation was identified in ICU charting (item identifiers with a positive amount) and in the electronic medication administration record (eMAR; medication names, administrations documented as given, with the documented dose) (Table S1). The eMAR administration was used when no basal insulin was charted in the ICU record; when both sources recorded an administration, the ICU-charted type and dose were retained and the eMAR time was used only when it was more than 30 min earlier. When both sources were available, doses were identical in 98.9% of episodes and times were within 30 min in 97.7% (Table S2). The IV total daily dose was 24 times the insulin infused during the final 6 h (rate multiplied by the duration of each record within the window) divided by 6, so that interruptions within the episode contributed zero rate.

**Outcomes and follow-up.** Each outcome was the first glucose value beyond its threshold after discontinuation. Follow-up ended at the end of the outcome window, ICU discharge or resumption of IV insulin, whichever came first; resumption was counted when the next infusion episode started within 48 h and before ICU discharge. Glucose values describing what was known at the bedside before discontinuation (last point-of-care value and values below 70 mg/dL in the final 6 h) were restricted to fingerstick and whole-blood measurements, because serum values become available only after sampling. Follow-up was limited to the ICU because point-of-care glucose values measured on the ward are not recorded in the database: among 11,720 ICU stays with an IV insulin infusion of any duration that were followed by at least 24 h on the ward, the median number of glucose values recorded in the laboratory database during the first 24 h after ICU discharge was one, and only 67 stays had four or more values.

**Covariates.** Definitions, time windows, functional forms and missing data are given in Table S3. Vasopressor dose was expressed in norepinephrine equivalents (µg/kg/min): norepinephrine + epinephrine + phenylephrine/10 + dopamine/100 + vasopressin (U/min) × 2.5 [26]. Diet was the most recent “Diet Type” entry within 24 h before discontinuation, grouped as nothing by mouth, liquids or sips, oral diet (any solid diet), enteral tube feeding, parenteral nutrition or not recorded. Insulin use before admission was ascertained from the “Medications on Admission” section of the discharge summary and from emergency department medication reconciliation, and was classified as present when either source mentioned insulin, absent when at least one source was available without such mention, and unknown otherwise; all 411 episodes in the 2020–2022 anchor year group were unknown because the note and emergency department modules do not include these admissions. Only binary indicators were extracted from the notes, and no free text left the database environment. Calendar period was the anchor year group, a de-identified 3-year interval.

**Insulin use after discontinuation.** Short-acting insulin comprised ICU-charted rapid-acting analogues, U-500 insulin and regular insulin boluses, and eMAR insulin administrations without an infusion rate and without a basal insulin name; an eMAR administration within 60 min of an ICU-charted administration of the same class was treated as a duplicate. Regular insulin boluses could not be classified as subcutaneous or IV. Short-acting insulin before hypoglycemia was any dose within 4 h before the first glucose value below 70 mg/dL, and additional basal insulin was any basal dose given more than 1 h after discontinuation during the 48-h follow-up (Tables S12 and S15).

**Cox models and multiple imputation.** Cox models used the Breslow method for ties and robust variance clustered by patient [27]. The estimation code was validated with simulated data and against an independent optimization routine, and unadjusted estimates were reproduced with the lifelines and statsmodels packages. In one imputed data set, the fully adjusted models were also fitted with the lifelines package (Efron method for ties): HRs per 0.1 increase in the ratio were identical to four decimal places (0.9578 for rebound hyperglycemia and 1.1158 for hypoglycemia), robust standard errors differed by less than 0.001, and the largest difference across all coefficients was 0.005. Continuous covariates were modeled with restricted cubic splines with three knots (Table S3), except vasopressor dose, whose 10th and 50th percentiles were both zero; compared with linear terms, this specification reduced the Akaike information criterion by 176 for rebound hyperglycemia and by 6 for hypoglycemia. Tests of the overall association and of nonlinearity for the ratio were multiparameter Wald tests pooled with the D1 statistic [S1]. Missing covariate values (Table S3) were imputed 20 times by chained equations with Bayesian ridge regression and posterior predictive draws (15 iterations, values bounded to plausible ranges); the imputation model included all covariates, the exposure, the event indicators and the Nelson–Aalen cumulative hazards of both primary outcomes [30], and estimates were combined with Rubin’s rules [31]. Unknown insulin use before admission was analyzed as a separate category.

**Proportional hazards, intervals and subgroups.** Proportional hazards were assessed with score tests based on scaled Schoenfeld residuals, and the median P value across imputations is reported [29]. Interval-specific estimates were obtained among episodes still at risk at the start of each interval (0–6, 6–12, 12–24 and 6–24 h), with time measured from the start of the interval (Table S10). Subgroup analyses added a product term between the ratio and each subgroup indicator to the fully adjusted model; the interaction with oral diet was also estimated separately for 0–6 and 6–24 h (Table S13).

**Timing of glargine.** Timing was categorized as after discontinuation, and 0 to <1, 1 to <2 (reference), 2 to <3 and ≥3 h before discontinuation. These models excluded variables that may be affected by an early dose (infusion rate over the final 6 h, rate trend, final infusion rate and glucose values in the final 6 h) and included instead the mean infusion rate 6–12 h before discontinuation and the glargine dose per kilogram; the comparison of doses given at least 2 h before with those given 0 to <2 h before discontinuation excluded doses given after discontinuation (Table S11).

**Standardized risks and shape of the dose–response relationship.** In each imputed data set, the fully adjusted models with the spline-modeled ratio were fitted, and each episode’s risk was predicted with its ratio set to selected values and averaged over the cohort (parametric g-formula), using the Breslow baseline cumulative hazard. For rebound hyperglycemia, for which hazards were not proportional, separate models were fitted for 0–6 h (all episodes) and 6–24 h (episodes still at risk at 6 h), and the 24-h risk was one minus the product of the two predicted survival probabilities; for hypoglycemia, a single model for 48 h was used. Variances were estimated from 50 patient-level bootstrap samples per imputed data set (1,000 in total) and combined with Rubin’s rules (Table 3 of the main text). The shape of the relationship was examined with three knots (10th, 50th and 90th percentiles) and five knots (5th, 27.5th, 50th, 72.5th and 95th percentiles), with the highest category divided at 0.80, and, for rebound hyperglycemia, among episodes still at risk 6 h after discontinuation (Table S9).

**Clinically significant hypoglycemia.** Because this outcome had 55 events, estimates were adjusted for five covariates (six parameters) entered as linear terms: diabetes, insulin use before admission (yes and unknown vs no), body weight, log creatinine and last point-of-care glucose, the strongest correlates of hypoglycemia in the primary model (Table S8).

**Sensitivity analyses.** The analyses are numbered as in Table S14: (1) complete-case analysis; (2) inverse probability of censoring weighting, in which the probability of remaining uncensored by ICU discharge or resumption of IV insulin was modeled in 1-h intervals with pooled logistic regression including all covariates, the exposure and a spline of time, and stabilized weights truncated at the 1st and 99th percentiles were applied in a pooled logistic outcome model whose odds ratios approximate hazard ratios [33]; (3) at least 24 h of potential ICU follow-up; (4) IV total daily dose from the insulin infused over up to 24 h, divided by the duration of the episode within that window; (5) first episode of each patient; (6) exclusion of glargine given after discontinuation; (7) exclusion of point-of-care glucose below 70 mg/dL in the final 6 h; (8) exclusion of basal insulin 12–48 h before discontinuation; (9) exposure and eligibility from ICU charting alone; (10) all basal insulin types, with type indicators; (11) exclusion of type 1 diabetes; (12) a parsimonious adjustment set (age, sex, diabetes, HbA1c, insulin use before admission, body weight, creatinine, mean infusion rate and glucose over the final 6 h, rate trend, oral diet, glargine at least 2 h before discontinuation and cardiac surgery); and (13) linear terms for continuous covariates.

Analyses (14) to (21) were: (14) exclusion of ratios of 10/24 (0.417 ± 0.0025), corresponding to a dose of 10 times the mean hourly infusion rate; (15) ratios of 1.0 or less; (16) exclusion of the 2020–2022 anchor year group; (17) hypoglycemia within 24 h; (18) censoring of hypoglycemia events preceded by short-acting insulin within 4 h; (19) the ratio, the mean infusion rate over the final 6 h and the rate trend recalculated over the time the infusion was running; (20) exclusion of episodes with an interruption of 15 min or longer within the final 6 h; and (21) exclusion of episodes with an interruption of 30 min or longer between the glargine dose and discontinuation. For analyses (19) to (21), interruptions, defined as intervals between consecutive charted infusion records within an episode, were extracted with a separate query using the same episode definition, which reproduced all 13,824 infusion episodes and matched all 6,220 analyzed episodes. The glargine dose per 0.1 U/kg was examined as an alternative exposure, and E-values were calculated from unrounded hazard ratios, treated as approximations of risk ratios because hypoglycemia was uncommon [34].

**Software.** Analyses were performed in Python 3.12.9 with NumPy, pandas, SciPy and scikit-learn.

**Table S1 Data elements, source tables and codes**

| **Data element** | **Source** | **Identifiers, codes or search terms** | **Use** |
| --- | --- | --- | --- |
| IV regular insulin infusion | icu.inputevents | Item 223258 (Insulin – Regular) with rate >0; records with status “Rewritten” excluded | Infusion episodes, exposure denominator, resumption |
| Basal insulin, ICU charting | icu.inputevents | Items 223260 (glargine), 223259 (NPH), 223257 (70/30), 223261 (Humalog 75/25) with amount >0 | Exposure; prior basal insulin; additional basal insulin |
| Basal insulin, eMAR | hosp.emar, hosp.emar_detail | Names: glargine, Lantus, Basaglar, Toujeo, Semglee (glargine); detemir, Levemir; NPH, isophane, Humulin N, Novolin N; premixed 70/30, 75/25, 50/50. Events: administered, delayed administered, partially administered, confirmed. Dose: dose given | Exposure reconciliation; prior basal insulin |
| Glucose | icu.chartevents | Items 225664 (fingerstick), 220621 (serum), 226537 (whole blood); >10 and <1,000 mg/dL | Outcomes; mean glucose before discontinuation |
| Point-of-care glucose before discontinuation | icu.chartevents | Items 225664 and 226537 | Last value; values <70 mg/dL in final 6 h |
| Short-acting insulin after discontinuation | icu.inputevents; hosp.emar, hosp.emar_detail | Items 223262 (Humalog), 229299 (Novolog), 229619 (U-500), and 223258 recorded as a bolus (Drug Push); eMAR insulin without infusion rate and without a basal name | Tables S12 and S15 |
| Vasopressors | icu.inputevents | Items 221906 (norepinephrine), 221289 (epinephrine), 221749, 229630, 229632 (phenylephrine), 221662 (dopamine), 222315 (vasopressin); rate >0 | Vasopressor dose |
| Enteral and parenteral nutrition | icu.inputevents, icu.d_items | Categories “Nutrition – Enteral” and “Nutrition – Parenteral”; lipid emulsions (225801, 227090) excluded; rate >0 | Nutrition indicator |
| Dextrose infusion | icu.inputevents | Items 220950 (10%), 228140 (20%), 228141 (30%), 228142 (40%); rate >0 | Covariate |
| Diet | icu.chartevents | Item 224001 (Diet Type) | Oral diet; nutritional status |
| Corticosteroids | hosp.prescriptions | Hydrocortisone, methylprednisolone, prednisone, prednisolone, dexamethasone; IV, intramuscular or enteral route | Covariate |
| HbA1c | hosp.labevents | Item 50852; 3–20% | Covariate |
| Creatinine | hosp.labevents | Item 50912 | Covariate |
| Weight and height | icu.inputevents; icu.chartevents | Patient weight on infusion records; “Admission Weight (Kg)” or “Daily Weight” (20–400 kg), used when the infusion-record weight was outside 30–300 kg; “Height (cm)” (100–250 cm) | Body weight; body mass index |
| Diabetes | hosp.diagnoses_icd | ICD-9-CM 250; ICD-10-CM E10, E11, E13 | Covariate |
| Type 1 diabetes | hosp.diagnoses_icd | ICD-9-CM 250.x1, 250.x3; ICD-10-CM E10 | Sensitivity analysis (11) |
| Diabetic ketoacidosis or hyperosmolar hyperglycemic state | hosp.diagnoses_icd | ICD-9-CM 250.1, 250.2; ICD-10-CM codes beginning with E10.1, E11.0, E11.1, E13.0 or E13.1 | Exclusion |
| Coronary artery bypass grafting | hosp.procedures_icd | ICD-9-CM 36.1; ICD-10-PCS 0210–0213; procedure date on or before discontinuation | Cardiac surgery |
| Valve surgery | hosp.procedures_icd | ICD-9-CM 35.1, 35.2; ICD-10-PCS 02Q, 02R or 02U with body part F, G, H or J | Cardiac surgery |
| Heart failure; chronic kidney disease | hosp.diagnoses_icd | ICD-9-CM 428, ICD-10-CM I50; ICD-9-CM 585, ICD-10-CM N18 | Table S4 |
| Invasive ventilation | icu.procedureevents | “Invasive Ventilation” | Table S4 |
| SOFA score and Charlson index | Derived tables (first_day_sofa, charlson) | MIMIC Code Repository concepts [25] | Covariates |
| Insulin use before admission | note.discharge; ed.medrecon, ed.edstays | Insulin, glargine, Lantus, Levemir, detemir, Basaglar, Toujeo, Semglee, Tresiba, degludec, Humalog, Novolog, lispro, aspart, Humulin, Novolin, Apidra, Admelog | Covariate |
| Non-insulin glucose-lowering drugs before admission | note.discharge; ed.medrecon, ed.edstays | Metformin, glipizide, glyburide, glimepiride, sitagliptin, Januvia, linagliptin, saxagliptin, alogliptin, pioglitazone, empagliflozin, Jardiance, dapagliflozin, Farxiga, canagliflozin, ertugliflozin, liraglutide, Victoza, semaglutide, Ozempic, Rybelsus, dulaglutide, Trulicity, exenatide, tirzepatide, Mounjaro, repaglinide, nateglinide, acarbose | Table S4 |

*Tables are from MIMIC-IV version 3.1 (hosp and icu modules), MIMIC-IV-ED version 2.2 (ed) and MIMIC-IV-Note version 2.2 (note). Name searches were case-insensitive. Valve procedure codes were not restricted by approach and may include transcatheter procedures. U-500 regular insulin was grouped with short-acting insulin. eMAR, electronic medication administration record; HbA1c, glycated hemoglobin; ICD-9-CM and ICD-10-CM, International Classification of Diseases, Ninth and Tenth Revisions, Clinical Modification; ICD-10-PCS, ICD-10 Procedure Coding System; ICU, intensive care unit; IV, intravenous; NPH, neutral protamine Hagedorn; SOFA, Sequential Organ Failure Assessment.*

**Table S2 Recording of glargine in ICU charting and the electronic medication administration record**

*A. Glargine administrations recorded in the eMAR during ICU stays in the source population*

| **Calendar period** | **eMAR administrations, n** | **Without a corresponding ICU-charted record, n (%)** |
| --- | --- | --- |
| 2008–2010 | 258 | 44 (17.1) |
| 2011–2013 | 213 | 32 (15.0) |
| 2014–2016 | 750 | 59 (7.9) |
| 2017–2019 | 1,367 | 70 (5.1) |
| 2020–2022 | 894 | 538 (60.2) |
| All periods | 3,482 | 743 (21.3) |

*B. Source of the first glargine dose among the 6,220 transition episodes*

| **Category** | **Episodes, n (%)** |
| --- | --- |
| First glargine dose recorded in ICU charting only | 3,669 (59.0) |
| First glargine dose recorded in both sources | 2,469 (39.7) |
| Identical dose in both sources | 2,441 (98.9) |
| Time difference (ICU charting minus eMAR), h, median (IQR) | 0.00 (−0.03 to 0.04) |
| Times within 30 min of each other | 2,412 (97.7) |
| eMAR time more than 30 min earlier (eMAR time used) | 17 (0.7) |
| eMAR time more than 30 min later (ICU-charted time retained) | 40 (1.6) |
| First glargine dose recorded in the eMAR only (eMAR type, dose and time used) | 82 (1.3) |
| By calendar period, n: 2008–2010 / 2011–2013 / 2014–2016 / 2017–2019 / 2020–2022 | 6 / 7 / 21 / 30 / 18 |

*In panel A, administrations recorded as administered or delayed administered during an ICU stay were compared with glargine records in ICU charting (source population: all ICU stays with IV insulin); partially administered and confirmed administrations, which were included in the exposure definition, did not occur for glargine during ICU stays. In panel B, percentages of the subcategories refer to episodes with records in both sources. Calendar period is the de-identified anchor year group. eMAR, electronic medication administration record; ICU, intensive care unit; IQR, interquartile range.*

**Table S3 Covariate definitions, time windows, model form and missing data**

| **Covariate** | **Definition and time window** | **Form in model** | **Missing, n (%)** |
| --- | --- | --- | --- |
| Age, years | Age at discontinuation | Restricted cubic spline (knots 52, 67, 80) | 0 |
| Sex | Recorded sex (male vs female) | Binary | 0 |
| Diabetes | Diagnosis code for diabetes during the admission (Table S1) | Binary | 0 |
| HbA1c, % | Most recent value from 90 days before admission to hospital discharge (plausible range 3–20%) | Restricted cubic spline (knots 5.3, 6.0, 8.4) | 402 (6.5) |
| Insulin use before admission | Insulin named in the "Medications on Admission" section of the discharge summary or in emergency department medication reconciliation; unknown when neither source was available | Categorical: yes and unknown vs no | 882 (14.2) unknown, analyzed as a category |
| Body weight, kg | Weight recorded on the infusion records of the episode; charted admission or daily weight when outside 30–300 kg | Restricted cubic spline (knots 65.2, 86.7, 113.0) | 0 |
| Creatinine, mg/dL | Most recent serum creatinine within 24 h before discontinuation; log-transformed (bounded to 0.2–15 mg/dL) | Restricted cubic spline (knots 0.60, 0.90, 1.60) | 5 (0.1) |
| Charlson comorbidity index | Derived admission-level index [22, 23, 25] | Restricted cubic spline (knots 2, 4, 8) | 0 |
| SOFA score, first ICU day | Derived score for the first 24 h of the ICU stay [24, 25] | Restricted cubic spline (knots 2, 5, 9) | 0 |
| Infusion duration, h | Time from the start to the end of the infusion episode; log-transformed | Restricted cubic spline (knots 10.0, 15.8, 29.0) | 0 |
| Mean infusion rate in final 6 h, U/h | Insulin infused during the final 6 h divided by 6; log-transformed | Restricted cubic spline (knots 1.00, 2.28, 4.76) | 0 |
| Rate trend | Mean rate in the final 6 h divided by the mean rate over the part of the period 6–12 h before discontinuation covered by the episode; truncated at 5 | Restricted cubic spline (knots 0.39, 0.81, 1.58) | 4 (0.1) |
| Final infusion rate, U/h | Rate of the last charted infusion record; log(1 + rate) | Restricted cubic spline (knots 1.0, 2.0, 4.0) | 0 |
| Mean glucose in final 6 h, mg/dL | Mean of all ICU glucose values in the 6 h before discontinuation | Restricted cubic spline (knots 102, 117, 143) | 1 (<0.1) |
| Last point-of-care glucose, mg/dL | Last fingerstick or whole-blood glucose value within 6 h before discontinuation | Restricted cubic spline (knots 89, 112, 145) | 22 (0.4) |
| Vasopressor dose, µg/kg/min | Norepinephrine-equivalent dose of vasopressor infusions running at discontinuation [26]; 0 if none | Linear (truncated at the 99th percentile, 0.18) | 0 |
| Enteral or parenteral nutrition | Enteral nutrition running at discontinuation or parenteral nutrition (excluding lipid emulsions) within 2 h before | Binary | 0 |
| Oral diet at discontinuation | Most recent "Diet Type" entry within 24 h before discontinuation indicating an oral diet | Binary | 0 |
| Dextrose infusion | Infusion of 10–40% dextrose within 2 h before discontinuation | Binary | 0 |
| Corticosteroid therapy | Systemic corticosteroid prescription active at discontinuation | Binary | 0 |
| Cardiac vascular ICU | First care unit of the ICU stay | Binary | 0 |
| Cardiac surgery before discontinuation | Coronary artery bypass grafting or valve surgery code with a procedure date on or before the date of discontinuation | Binary | 0 |
| ICU time before discontinuation, h | Time from ICU admission to discontinuation; log(1 + h) | Restricted cubic spline (knots 17.1, 21.1, 67.5) | 0 |
| Basal insulin 12–48 h before discontinuation | Any basal insulin in ICU charting or the eMAR 12–48 h before discontinuation | Binary | 0 |
| Glargine ≥2 h before discontinuation | Reconciled time of the first glargine dose at least 2 h before discontinuation | Binary | 0 |
| Calendar period | Anchor year group: 2008–2010 (reference), 2011–2013, 2014–2016, 2017–2019, 2020–2022 | Categorical | 0 |

*All covariates were included in the adjusted models. HbA1c values measured after discontinuation were eligible because HbA1c reflects glycemia over the preceding 2–3 months. Spline knots were placed at the 10th, 50th and 90th percentiles of the observed values in the cohort and are shown on the original scale. Missing values were imputed (Supplementary Methods). eMAR, electronic medication administration record; HbA1c, glycated hemoglobin; ICU, intensive care unit; SOFA, Sequential Organ Failure Assessment.*

**Table S4 Complete characteristics of transition episodes by glargine-to-intravenous dose ratio**

| **Characteristic** | **Overall** | **<0.30** | **0.30–0.39** | **0.40–0.49 (reference)** | **0.50–0.59** | **≥0.60** |
| --- | --- | --- | --- | --- | --- | --- |
| Episodes, n | 6,220 | 1,528 | 1,473 | 2,104 | 548 | 567 |
| Patients, n | 5,924 | 1,474 | 1,461 | 2,086 | 546 | 556 |
| **Demographics and comorbidity** |  |  |  |  |  |  |
| Age, years | 67 (60–74) | 67 (60–74) | 68 (60–74) | 67 (60–74) | 68 (60–75) | 66 (60–75) |
| Male sex | 4,409 (70.9) | 1,030 (67.4) | 1,073 (72.8) | 1,563 (74.3) | 384 (70.1) | 359 (63.3) |
| Race: White | 4,510 (72.5) | 1,082 (70.8) | 1,094 (74.3) | 1,569 (74.6) | 379 (69.2) | 386 (68.1) |
| Race: Black | 296 (4.8) | 98 (6.4) | 59 (4.0) | 75 (3.6) | 26 (4.7) | 38 (6.7) |
| Race: other | 589 (9.5) | 138 (9.0) | 133 (9.0) | 185 (8.8) | 57 (10.4) | 76 (13.4) |
| Race: unknown | 825 (13.3) | 210 (13.7) | 187 (12.7) | 275 (13.1) | 86 (15.7) | 67 (11.8) |
| Body mass index, kg/m² | 29.2 (26.0–33.3) | 29.4 (26.0–33.4) | 29.6 (26.4–34.3) | 29.2 (26.2–33.0) | 28.5 (25.7–32.3) | 28.4 (24.8–32.4) |
| Weight, kg | 86.7 (75.0–100.0) | 86.9 (75.0–100.0) | 88.0 (77.0–102.0) | 87.8 (76.0–100.0) | 85.4 (74.8–97.5) | 82.5 (70.0–96.2) |
| Diabetes (ICD code) | 2,861 (46.0) | 825 (54.0) | 657 (44.6) | 799 (38.0) | 252 (46.0) | 328 (57.8) |
| Type 1 diabetes | 174 (2.8) | 63 (4.1) | 32 (2.2) | 21 (1.0) | 19 (3.5) | 39 (6.9) |
| HbA1c, % | 6.0 (5.6–7.1) | 6.1 (5.6–7.5) | 6.0 (5.6–7.0) | 5.9 (5.5–6.7) | 6.0 (5.5–7.1) | 6.4 (5.7–7.7) |
| HbA1c ≥8.0% | 784 (12.6) | 237 (15.5) | 182 (12.4) | 186 (8.8) | 66 (12.0) | 113 (19.9) |
| HbA1c missing | 402 (6.5) | 165 (10.8) | 65 (4.4) | 86 (4.1) | 35 (6.4) | 51 (9.0) |
| Insulin use before admission: yes | 1,053 (16.9) | 330 (21.6) | 223 (15.1) | 245 (11.6) | 97 (17.7) | 158 (27.9) |
| Insulin use before admission: no | 4,285 (68.9) | 971 (63.5) | 1,040 (70.6) | 1,564 (74.3) | 383 (69.9) | 327 (57.7) |
| Insulin use before admission: unknown | 882 (14.2) | 227 (14.9) | 210 (14.3) | 295 (14.0) | 68 (12.4) | 82 (14.5) |
| Non-insulin glucose-lowering drug before admission | 1,533 (24.6) | 419 (27.4) | 351 (23.8) | 467 (22.2) | 144 (26.3) | 152 (26.8) |
| Chronic kidney disease | 1,117 (18.0) | 336 (22.0) | 251 (17.0) | 312 (14.8) | 89 (16.2) | 129 (22.8) |
| Heart failure | 1,584 (25.5) | 434 (28.4) | 347 (23.6) | 523 (24.9) | 150 (27.4) | 130 (22.9) |
| Charlson comorbidity index | 4 (3–6) | 4 (3–6) | 4 (3–6) | 4 (3–5) | 4 (3–6) | 4 (3–6) |
| **Admission and ICU** |  |  |  |  |  |  |
| Admission type: elective or same-day surgical | 2,838 (45.6) | 606 (39.7) | 697 (47.3) | 1,054 (50.1) | 246 (44.9) | 235 (41.4) |
| Admission type: urgent | 1,584 (25.5) | 402 (26.3) | 374 (25.4) | 513 (24.4) | 144 (26.3) | 151 (26.6) |
| Admission type: emergency | 1,173 (18.9) | 342 (22.4) | 253 (17.2) | 347 (16.5) | 117 (21.4) | 114 (20.1) |
| Admission type: observation or other | 625 (10.0) | 178 (11.6) | 149 (10.1) | 190 (9.0) | 41 (7.5) | 67 (11.8) |
| Cardiac surgery before discontinuation | 5,440 (87.5) | 1,201 (78.6) | 1,345 (91.3) | 1,948 (92.6) | 480 (87.6) | 466 (82.2) |
| Coronary artery bypass grafting | 3,955 (63.6) | 898 (58.8) | 993 (67.4) | 1,383 (65.7) | 342 (62.4) | 339 (59.8) |
| Valve surgery | 2,332 (37.5) | 532 (34.8) | 547 (37.1) | 837 (39.8) | 212 (38.7) | 204 (36.0) |
| Cardiac vascular ICU | 5,753 (92.5) | 1,280 (83.8) | 1,409 (95.7) | 2,050 (97.4) | 514 (93.8) | 500 (88.2) |
| SOFA score, first ICU day | 5 (3–7) | 5 (4–7) | 5 (3–7) | 5 (4–7) | 5 (3–7) | 5 (3–7) |
| Invasive ventilation during ICU stay | 5,862 (94.2) | 1,386 (90.7) | 1,398 (94.9) | 2,025 (96.2) | 523 (95.4) | 530 (93.5) |
| ICU time before discontinuation, h | 21.1 (19.5–24.7) | 21.2 (18.8–33.7) | 21.0 (19.4–23.5) | 21.1 (19.7–23.0) | 21.0 (19.7–24.4) | 21.3 (19.5–35.5) |
| Creatinine before discontinuation, mg/dL | 0.90 (0.70–1.20) | 1.00 (0.80–1.30) | 0.90 (0.80–1.20) | 0.90 (0.70–1.10) | 0.90 (0.78–1.20) | 0.90 (0.70–1.20) |
| **Intravenous insulin before discontinuation** |  |  |  |  |  |  |
| Infusion duration, h | 15.8 (12.8–18.4) | 15.5 (12.0–19.1) | 15.6 (12.7–18.2) | 16.0 (13.3–18.2) | 16.0 (13.3–18.6) | 15.6 (13.1–18.6) |
| Mean rate in final 6 h, U/h | 2.28 (1.59–3.36) | 3.04 (1.98–4.54) | 2.51 (1.60–3.67) | 2.00 (1.68–3.00) | 2.13 (1.53–2.89) | 1.36 (1.00–2.06) |
| Estimated IV total daily dose, U | 54.8 (38.1–80.7) | 73.0 (47.5–108.9) | 60.1 (38.4–88.0) | 48.0 (40.4–72.0) | 51.2 (36.7–69.3) | 32.6 (24.0–49.5) |
| Rate trend (final 6 h / preceding 6 h) | 0.81 (0.56–1.11) | 0.90 (0.64–1.30) | 0.82 (0.58–1.09) | 0.82 (0.59–1.07) | 0.73 (0.47–1.02) | 0.54 (0.36–0.83) |
| Final infusion rate, U/h | 2.0 (1.0–3.0) | 2.0 (1.0–3.0) | 2.0 (2.0–3.0) | 2.0 (1.0–3.0) | 2.0 (2.0–3.0) | 2.0 (1.0–3.0) |
| Mean glucose in final 6 h, mg/dL | 117 (108–127) | 117 (107–132) | 115 (107–124) | 115 (108–124) | 120 (113–128) | 122 (114–134) |
| Last point-of-care glucose, mg/dL | 112 (100–127) | 112 (97–128) | 112 (101–126) | 112 (100–124) | 115 (102–128) | 119 (103–136) |
| Point-of-care glucose <70 mg/dL in final 6 h | 102 (1.6) | 27 (1.8) | 17 (1.2) | 22 (1.0) | 14 (2.6) | 22 (3.9) |
| **First glargine dose** |  |  |  |  |  |  |
| Dose, U | 20 (10–30) | 16 (10–20) | 20 (15–30) | 20 (20–30) | 30 (20–40) | 30 (20–40) |
| Dose, U/kg | 0.24 (0.16–0.34) | 0.17 (0.12–0.25) | 0.24 (0.17–0.34) | 0.25 (0.18–0.34) | 0.32 (0.24–0.41) | 0.32 (0.23–0.47) |
| Glargine-to-IV dose ratio | 0.41 (0.30–0.46) | 0.23 (0.18–0.27) | 0.35 (0.33–0.38) | 0.42 (0.42–0.45) | 0.54 (0.52–0.56) | 0.72 (0.64–0.85) |
| Timing relative to discontinuation, h | −1.8 (−2.0 to −1.1) | −1.7 (−2.0 to −0.9) | −1.8 (−2.0 to −1.3) | −1.8 (−2.0 to −1.4) | −1.7 (−2.0 to −1.2) | −1.7 (−2.0 to −1.0) |
| Given ≥2 h before discontinuation | 1,890 (30.4) | 429 (28.1) | 447 (30.3) | 704 (33.5) | 154 (28.1) | 156 (27.5) |
| Given after discontinuation | 355 (5.7) | 159 (10.4) | 58 (3.9) | 75 (3.6) | 23 (4.2) | 40 (7.1) |
| Basal insulin 12–48 h before discontinuation | 355 (5.7) | 123 (8.0) | 56 (3.8) | 58 (2.8) | 31 (5.7) | 87 (15.3) |
| Exposure recorded only in eMAR | 82 (1.3) | 23 (1.5) | 14 (1.0) | 28 (1.3) | 9 (1.6) | 8 (1.4) |
| **Status at discontinuation** |  |  |  |  |  |  |
| Diet: nothing by mouth | 2,447 (39.3) | 675 (44.2) | 570 (38.7) | 798 (37.9) | 201 (36.7) | 203 (35.8) |
| Diet: liquids or sips | 2,723 (43.8) | 518 (33.9) | 690 (46.8) | 1,040 (49.4) | 256 (46.7) | 219 (38.6) |
| Diet: oral diet | 738 (11.9) | 197 (12.9) | 155 (10.5) | 201 (9.6) | 67 (12.2) | 118 (20.8) |
| Diet: enteral tube feeding | 249 (4.0) | 110 (7.2) | 45 (3.1) | 46 (2.2) | 21 (3.8) | 27 (4.8) |
| Diet: parenteral nutrition | 11 (0.2) | 5 (0.3) | 3 (0.2) | 3 (0.1) | 0 (0.0) | 0 (0.0) |
| Diet: not recorded | 52 (0.8) | 23 (1.5) | 10 (0.7) | 16 (0.8) | 3 (0.5) | 0 (0.0) |
| Enteral or parenteral nutrition infusion | 317 (5.1) | 150 (9.8) | 56 (3.8) | 52 (2.5) | 25 (4.6) | 34 (6.0) |
| Dextrose infusion within 2 h | 30 (0.5) | 15 (1.0) | 4 (0.3) | 3 (0.1) | 3 (0.5) | 5 (0.9) |
| Corticosteroid therapy | 230 (3.7) | 110 (7.2) | 33 (2.2) | 39 (1.9) | 14 (2.6) | 34 (6.0) |
| Vasopressor infusion at discontinuation | 1,338 (21.5) | 327 (21.4) | 317 (21.5) | 431 (20.5) | 133 (24.3) | 130 (22.9) |
| **Calendar period** |  |  |  |  |  |  |
| 2008–2010 | 1,535 (24.7) | 383 (25.1) | 344 (23.4) | 533 (25.3) | 131 (23.9) | 144 (25.4) |
| 2011–2013 | 1,319 (21.2) | 313 (20.5) | 317 (21.5) | 446 (21.2) | 120 (21.9) | 123 (21.7) |
| 2014–2016 | 1,540 (24.8) | 379 (24.8) | 362 (24.6) | 516 (24.5) | 149 (27.2) | 134 (23.6) |
| 2017–2019 | 1,415 (22.7) | 351 (23.0) | 343 (23.3) | 476 (22.6) | 115 (21.0) | 130 (22.9) |
| 2020–2022 | 411 (6.6) | 102 (6.7) | 107 (7.3) | 133 (6.3) | 33 (6.0) | 36 (6.3) |

*Values are median (interquartile range) or number (%). Patients could contribute episodes to more than one ratio category. Body mass index was missing for 374 episodes and HbA1c for 402. Race and admission type were taken from the hospital admission record; elective or same-day surgical admissions include surgical same-day admissions, and emergency admissions include emergency ward and direct emergency admissions. Timing is expressed relative to discontinuation (negative values denote administration before discontinuation). The estimated IV total daily dose is 24 times the mean infusion rate over the final 6 h. Coronary artery bypass grafting and valve surgery were combined in 847 episodes. Diet refers to the most recent Diet Type entry within 24 h before discontinuation; enteral or parenteral nutrition infusion refers to infusions recorded in ICU charting. Non-insulin glucose-lowering drugs were ascertained from the same sources as insulin (Table S1), and status was unknown for the same 882 episodes. Calendar period is the de-identified anchor year group. eMAR, electronic medication administration record; HbA1c, glycated hemoglobin; ICU, intensive care unit; IV, intravenous; SOFA, Sequential Organ Failure Assessment.*

**Table S5 Distribution of the glargine-to-intravenous dose ratio**

*A. Percentiles*

| **Statistic** | **Ratio** |
| --- | --- |
| Minimum | 0.02 |
| 2.5th percentile | 0.12 |
| 5th percentile | 0.16 |
| 10th percentile | 0.21 |
| 25th percentile | 0.30 |
| Median | 0.41 |
| 75th percentile | 0.46 |
| 90th percentile | 0.58 |
| 95th percentile | 0.70 |
| 97.5th percentile | 0.83 |
| Maximum | 2.50 |

*B. Episodes by ratio interval*

| **Ratio interval** | **Episodes, n (%)** |
| --- | --- |
| <0.10 | 96 (1.5) |
| 0.10–0.19 | 440 (7.1) |
| 0.20–0.29 | 992 (15.9) |
| 0.30–0.39 | 1,473 (23.7) |
| 0.40–0.49 | 2,104 (33.8) |
| 0.50–0.59 | 548 (8.8) |
| 0.60–0.69 | 257 (4.1) |
| 0.70–0.79 | 107 (1.7) |
| 0.80–0.99 | 127 (2.0) |
| 1.00–1.49 | 59 (0.9) |
| ≥1.50 | 17 (0.3) |

*The ratio is the first glargine dose divided by 24 times the mean intravenous insulin infusion rate over the final 6 h before discontinuation (6,220 episodes). A ratio of 10/24 (0.417 ± 0.0025), corresponding to a dose of 10 times the mean hourly infusion rate, occurred in 1,048 episodes (16.8%). Intervals include the lower and exclude the upper boundary; the five categories used in the analyses combine these intervals (<0.30, 0.30–0.39, 0.40–0.49, 0.50–0.59 and ≥0.60).*

**Table S6 Cumulative incidence of the primary outcomes over time by glargine-to-intravenous dose ratio**

| **Outcome and time** | **Overall** | **<0.30** | **0.30–0.39** | **0.40–0.49 (reference)** | **0.50–0.59** | **≥0.60** |
| --- | --- | --- | --- | --- | --- | --- |
| Episodes, n | 6,220 | 1,528 | 1,473 | 2,104 | 548 | 567 |
| **Rebound hyperglycemia (>180 mg/dL)** |  |  |  |  |  |  |
| At 6 h, % (95% CI) | 20.3 (19.3–21.3) | 26.7 (24.5–29.0) | 20.8 (18.8–23.0) | 14.7 (13.2–16.3) | 16.7 (13.8–20.2) | 25.5 (22.1–29.4) |
| Number at risk after 6 h | 4,411 | 1,012 | 1,031 | 1,591 | 391 | 386 |
| At 12 h, % (95% CI) | 31.2 (30.0–32.5) | 41.3 (38.6–44.0) | 30.7 (28.2–33.4) | 23.5 (21.5–25.5) | 27.1 (23.3–31.5) | 36.9 (32.7–41.3) |
| Number at risk after 12 h | 2,413 | 550 | 559 | 869 | 223 | 212 |
| At 24 h, % (95% CI) | 44.4 (42.8–45.9) | 55.2 (52.2–58.3) | 43.6 (40.4–46.9) | 35.4 (32.8–38.1) | 44.8 (39.5–50.5) | 48.2 (43.3–53.3) |
| **Hypoglycemia (<70 mg/dL)** |  |  |  |  |  |  |
| At 12 h, % (95% CI) | 1.9 (1.6–2.3) | 2.0 (1.4–2.9) | 1.1 (0.6–1.8) | 1.6 (1.2–2.3) | 2.6 (1.4–4.5) | 4.2 (2.8–6.4) |
| Number at risk after 12 h | 3,498 | 939 | 800 | 1,126 | 309 | 324 |
| At 24 h, % (95% CI) | 3.4 (2.9–4.0) | 3.7 (2.7–5.0) | 2.1 (1.3–3.2) | 2.7 (2.0–3.8) | 4.4 (2.7–7.1) | 7.5 (5.3–10.6) |
| Number at risk after 24 h | 2,643 | 699 | 608 | 864 | 223 | 249 |
| At 48 h, % (95% CI) | 6.4 (5.5–7.4) | 6.4 (4.8–8.5) | 5.4 (3.7–7.8) | 5.8 (4.3–7.7) | 7.1 (4.5–11.0) | 10.6 (7.6–14.8) |

*Cumulative incidence was estimated with the Kaplan–Meier method, with censoring at ICU discharge or resumption of intravenous insulin; values at 24 h (rebound hyperglycemia) and 48 h (hypoglycemia) correspond to Table 2 of the main text. Numbers at risk are episodes still under observation without the outcome after the stated time. CI, confidence interval; ICU, intensive care unit.*

**Table S7 Adjusted hazard ratios for all covariates in the primary models**

| **Covariate** | **Contrast** | **Rebound hyperglycemia, HR (95% CI)** | **P†** | **Hypoglycemia, HR (95% CI)** | **P†** |
| --- | --- | --- | --- | --- | --- |
| Glargine-to-IV dose ratio, per 0.1 | Per 0.1 increase | 0.96 (0.93–0.99) | 0.006 | 1.11 (1.04–1.18) | 0.001 |
| Age, years | 74 vs 60 | 0.99 (0.91–1.08) | 0.98 | 1.07 (0.84–1.37) | 0.39 |
| Male sex |  | 1.05 (0.93–1.17) | 0.44 | 1.19 (0.86–1.66) | 0.29 |
| Diabetes |  | 2.12 (1.78–2.52) | <0.001 | 1.85 (1.17–2.92) | 0.008 |
| HbA1c, % | 7.1 vs 5.6 | 2.62 (2.15–3.19) | <0.001 | 1.00 (0.66–1.52) | 0.72 |
| Insulin use before admission: yes vs no |  | 1.15 (1.02–1.30) | 0.027 | 1.76 (1.17–2.65) | 0.006 |
| Insulin use before admission: unknown vs no |  | 1.09 (0.92–1.29) | 0.31 | 1.29 (0.75–2.19) | 0.35 |
| Joint test |  |  | 0.083 |  | 0.024 |
| Body weight, kg | 100 vs 75 | 0.96 (0.89–1.04) | 0.55 | 0.53 (0.44–0.65) | <0.001 |
| Creatinine, mg/dL | 1.2 vs 0.7 | 1.11 (1.01–1.23) | 0.011 | 1.49 (1.12–1.99) | 0.023 |
| Charlson comorbidity index | 6 vs 3 | 1.127 (1.004–1.264) | 0.13 | 0.88 (0.61–1.29) | 0.66 |
| SOFA score, first ICU day | 7 vs 3 | 0.92 (0.84–1.01) | 0.14 | 1.03 (0.78–1.36) | 0.32 |
| Infusion duration, h | 18.4 vs 12.8 | 1.20 (1.13–1.27) | <0.001 | 0.89 (0.76–1.04) | 0.27 |
| Mean infusion rate in final 6 h, U/h | 3.36 vs 1.59 | 1.41 (1.25–1.58) | <0.001 | 1.01 (0.73–1.40) | 0.18 |
| Rate trend | 1.11 vs 0.56 | 0.72 (0.65–0.79) | <0.001 | 1.14 (0.86–1.53) | 0.62 |
| Final infusion rate, U/h | 3.0 vs 1.0 | 0.91 (0.77–1.06) | 0.021 | 0.92 (0.60–1.41) | 0.93 |
| Mean glucose in final 6 h, mg/dL | 127 vs 108 | 1.33 (1.22–1.45) | <0.001 | 1.13 (0.88–1.44) | 0.027 |
| Last point-of-care glucose, mg/dL | 127 vs 100 | 1.13 (1.06–1.21) | <0.001 | 0.61 (0.49–0.75) | <0.001 |
| Vasopressor dose, µg/kg/min | 0.14 vs 0.00 | 1.25 (1.05–1.49) | 0.012 | 0.70 (0.38–1.30) | 0.26 |
| Enteral or parenteral nutrition |  | 1.62 (1.26–2.09) | <0.001 | 0.69 (0.33–1.44) | 0.32 |
| Oral diet at discontinuation |  | 1.49 (1.30–1.71) | <0.001 | 0.89 (0.56–1.43) | 0.64 |
| Dextrose infusion within 2 h |  | 0.98 (0.48–2.03) | 0.96 | 1.03 (0.22–4.73) | 0.97 |
| Corticosteroid therapy |  | 1.16 (0.92–1.47) | 0.20 | 1.02 (0.56–1.87) | 0.95 |
| Cardiac vascular ICU |  | 0.69 (0.52–0.92) | 0.012 | 1.63 (0.76–3.51) | 0.21 |
| Cardiac surgery before discontinuation |  | 1.06 (0.84–1.34) | 0.61 | 0.69 (0.40–1.22) | 0.20 |
| ICU time before discontinuation, h | 24.7 vs 19.5 | 0.92 (0.86–0.98) | 0.032 | 0.99 (0.84–1.17) | 0.56 |
| Basal insulin 12–48 h before discontinuation |  | 1.02 (0.84–1.23) | 0.85 | 1.77 (0.99–3.16) | 0.053 |
| Glargine ≥2 h before discontinuation |  | 0.79 (0.71–0.87) | <0.001 | 1.19 (0.89–1.60) | 0.25 |
| Calendar period 2011–2013 vs 2008–2010 |  | 1.07 (0.93–1.24) | 0.32 | 0.75 (0.52–1.09) | 0.13 |
| Calendar period 2014–2016 vs 2008–2010 |  | 1.21 (1.06–1.38) | 0.005 | 0.60 (0.40–0.88) | 0.009 |
| Calendar period 2017–2019 vs 2008–2010 |  | 1.21 (1.05–1.38) | 0.008 | 0.47 (0.30–0.74) | 0.001 |
| Calendar period 2020–2022 vs 2008–2010 |  | 1.12 (0.87–1.44) | 0.39 | 0.25 (0.08–0.77) | 0.015 |
| Joint test for calendar period |  |  | 0.028 |  | 0.002 |

*Estimates are from the fully adjusted Cox models for rebound hyperglycemia (glucose >180 mg/dL within 24 h; 2,109 events) and hypoglycemia (glucose <70 mg/dL within 48 h; 211 events) in 6,220 episodes, pooled over 20 imputations with robust variance clustered by patient. For continuous covariates, hazard ratios compare the 75th with the 25th percentile of observed values (vasopressor dose: 90th percentile among episodes receiving vasopressors vs none). †For covariates modeled with restricted cubic splines, the P value tests both spline terms jointly. Joint P values are shown for insulin use before admission and calendar period. Covariate estimates are mutually adjusted and should not be interpreted as causal effects. CI, confidence interval; HbA1c, glycated hemoglobin; HR, hazard ratio; ICU, intensive care unit; IV, intravenous; SOFA, Sequential Organ Failure Assessment.*

**Table S8 Secondary outcomes by glargine-to-intravenous dose ratio**

*A. Outcomes by ratio category and per 0.1 increase*

| **Outcome and group** | **Events/episodes** | **Cumulative incidence, % (95% CI)** | **Unadjusted HR (95% CI)** | **Adjusted HR (95% CI)** |
| --- | --- | --- | --- | --- |
| **Severe hyperglycemia (>250 mg/dL) within 24 h** |  |  |  |  |
| <0.30 | 269/1,528 | 23.3 (20.9–26.0) | 3.09 (2.49–3.84) | 1.16 (0.89–1.51) |
| 0.30–0.39 | 131/1,473 | 13.1 (11.0–15.5) | 1.55 (1.22–1.98) | 1.01 (0.78–1.30) |
| 0.40–0.49 | 122/2,104 | 8.6 (7.2–10.3) | 1.00 (reference) | 1.00 (reference) |
| 0.50–0.59 | 51/548 | 14.0 (10.7–18.4) | 1.63 (1.18–2.27) | 0.87 (0.61–1.25) |
| ≥0.60 | 65/567 | 15.1 (11.9–19.1) | 1.95 (1.43–2.65) | 0.77 (0.53–1.10) |
| Per 0.1 increase in ratio |  |  | 0.85 (0.78–0.92) | 0.93 (0.89–0.98) |
| **Resumption of IV insulin within 48 h** |  |  |  |  |
| <0.30 | 126/1,528 | 14.4 (12.1–17.0) | 1.93 (1.46–2.55) | 1.18 (0.86–1.62) |
| 0.30–0.39 | 73/1,473 | 9.7 (7.7–12.3) | 1.30 (0.95–1.77) | 1.06 (0.77–1.46) |
| 0.40–0.49 | 80/2,104 | 7.9 (6.3–9.9) | 1.00 (reference) | 1.00 (reference) |
| 0.50–0.59 | 28/548 | 9.9 (6.8–14.3) | 1.30 (0.85–2.00) | 0.96 (0.61–1.51) |
| ≥0.60 | 22/567 | 7.3 (4.7–11.0) | 0.93 (0.57–1.53) | 0.61 (0.37–1.02) |
| Per 0.1 increase in ratio |  |  | 0.85 (0.78–0.93) | 0.90 (0.83–0.98) |
| **Clinically significant hypoglycemia (<54 mg/dL) within 48 h** |  |  |  | Minimally adjusted HR (95% CI) |
| <0.30 | 17/1,528 | 1.9 (1.1–3.3) | 1.31 (0.67–2.55) | 1.05 (0.54–2.03) |
| 0.30–0.39 | 7/1,473 | 1.1 (0.5–2.3) | 0.62 (0.26–1.51) | 0.59 (0.24–1.44) |
| 0.40–0.49 | 16/2,104 | 1.8 (1.0–3.0) | 1.00 (reference) | 1.00 (reference) |
| 0.50–0.59 | 5/548 | 2.7 (1.1–6.7) | 1.15 (0.42–3.12) | 1.00 (0.37–2.69) |
| ≥0.60 | 10/567 | 3.5 (1.8–6.8) | 2.14 (0.97–4.71) | 1.50 (0.67–3.36) |
| Per 0.1 increase in ratio |  |  | 1.105 (1.002–1.219) | 1.08 (0.99–1.19) |

*B. Adjusted hazard ratios at selected ratios (restricted cubic splines)*

| **Glargine-to-IV dose ratio** | **Severe hyperglycemia** | **Resumption of IV insulin** | **Clinically significant hypoglycemia (minimally adjusted)** |
| --- | --- | --- | --- |
| 0.20 | 1.217 (1.001–1.479) | 1.36 (1.08–1.71) | 0.94 (0.57–1.57) |
| 0.30 | 1.06 (0.93–1.21) | 1.12 (0.95–1.33) | 0.92 (0.65–1.29) |
| 0.40 (reference) | 1.00 | 1.00 | 1.00 |
| 0.50 | 0.97 (0.86–1.09) | 0.93 (0.79–1.10) | 1.13 (0.77–1.66) |
| 0.60 | 0.92 (0.78–1.08) | 0.87 (0.70–1.10) | 1.25 (0.75–2.07) |
| 0.70 | 0.87 (0.72–1.06) | 0.82 (0.61–1.11) | 1.36 (0.81–2.28) |
| 0.80 | 0.82 (0.64–1.06) | 0.78 (0.52–1.17) | 1.49 (0.88–2.52) |
| P for overall association | 0.057 | 0.023 | 0.28 |
| P for nonlinearity | 0.59 | 0.48 | 0.90 |

*Cumulative incidence was estimated with the Kaplan–Meier method at 24 h (severe hyperglycemia) or 48 h (other outcomes). Severe hyperglycemia and resumption of intravenous insulin were analyzed with the fully adjusted models; clinically significant hypoglycemia (55 events) was adjusted for five covariates (Supplementary Methods). In panel B, the ratio was modeled with restricted cubic splines (knots at 0.16, 0.35, 0.42 and 0.70); hazard ratios are relative to a ratio of 0.40. Estimates were pooled over 20 imputations and are shown to three decimal places when a confidence limit rounds to 1.00. CI, confidence interval; HR, hazard ratio; IV, intravenous.*

**Table S9 Shape of the association between the glargine-to-intravenous dose ratio and the primary outcomes**

*A. Restricted cubic splines with different numbers of knots: hazard ratios (95% CI) between selected ratios*

| **Outcome and specification** | **Knots** | **0.30 vs 0.40** | **0.60 vs 0.40** | **0.80 vs 0.40** | **0.80 vs 0.60** | **P overall** | **P nonlinearity** |
| --- | --- | --- | --- | --- | --- | --- | --- |
| **Rebound hyperglycemia** |  |  |  |  |  |  |  |
| Three knots | 0.21, 0.41, 0.58 | 1.07 (1.01–1.12) | 0.93 (0.87–0.99) | 0.877 (0.767–1.004) | 0.94 (0.88–1.02) | 0.012 | 0.30 |
| Four knots (primary) | 0.16, 0.35, 0.42, 0.70 | 1.11 (1.04–1.19) | 0.87 (0.79–0.96) | 0.85 (0.75–0.97) | 0.98 (0.90–1.06) | 0.008 | 0.12 |
| Five knots | 0.16, 0.31, 0.41, 0.44, 0.70 | 1.04 (0.92–1.18) | 0.80 (0.68–0.94) | 0.79 (0.67–0.94) | 0.99 (0.92–1.07) | 0.011 | 0.11 |
| **Hypoglycemia** |  |  |  |  |  |  |  |
| Three knots | 0.21, 0.41, 0.58 | 0.83 (0.71–0.97) | 1.23 (1.07–1.41) | 1.45 (1.09–1.94) | 1.179 (1.004–1.385) | 0.006 | 0.28 |
| Four knots (primary) | 0.16, 0.35, 0.42, 0.70 | 0.81 (0.66–0.99) | 1.318 (1.004–1.729) | 1.52 (1.09–2.13) | 1.16 (0.96–1.39) | 0.016 | 0.47 |
| Five knots | 0.16, 0.31, 0.41, 0.44, 0.70 | 0.90 (0.60–1.36) | 1.47 (0.92–2.34) | 1.67 (1.05–2.67) | 1.14 (0.93–1.39) | 0.029 | 0.60 |

*B. Six ratio categories*

| **Glargine-to-IV dose ratio** | **Episodes, n** | **Rebound events** | **Rebound hyperglycemia, HR (95% CI)** | **Hypoglycemia events** | **Hypoglycemia, HR (95% CI)** |
| --- | --- | --- | --- | --- | --- |
| <0.30 | 1,528 | 681 | 1.12 (0.98–1.29) | 57 | 0.83 (0.53–1.30) |
| 0.30–0.39 | 1,473 | 487 | 1.13 (0.99–1.28) | 36 | 0.84 (0.54–1.31) |
| 0.40–0.49 (reference) | 2,104 | 545 | 1.00 | 58 | 1.00 |
| 0.50–0.59 | 548 | 174 | 0.86 (0.71–1.04) | 22 | 1.25 (0.77–2.04) |
| 0.60–0.79 | 364 | 136 | 0.87 (0.70–1.07) | 23 | 1.75 (1.03–2.98) |
| ≥0.80 | 203 | 86 | 0.82 (0.62–1.08) | 15 | 1.77 (0.86–3.66) |

*C. Four-knot spline among episodes at risk 6 h after discontinuation*

| **Comparison** | **Rebound hyperglycemia between 6 and 24 h, HR (95% CI)** |
| --- | --- |
| 0.30 vs 0.40 | 1.13 (1.02–1.26) |
| 0.60 vs 0.40 | 0.88 (0.77–1.01) |
| 0.80 vs 0.40 | 0.78 (0.62–0.98) |
| 0.80 vs 0.60 | 0.89 (0.76–1.04) |
| P for overall association | <0.001 |
| P for nonlinearity | 0.061 |

*Estimates are from fully adjusted Cox models pooled over 20 imputations. Knots were placed at the 10th, 50th and 90th percentiles (three knots), the 5th, 35th, 65th and 95th percentiles (four knots) or the 5th, 27.5th, 50th, 72.5th and 95th percentiles (five knots) of the ratio. P values are multiparameter Wald tests pooled with the D1 statistic. Estimates in panel B are from a separate six-category model and may differ slightly from those in Table 2 of the main text. Panel C includes episodes still at risk of rebound hyperglycemia 6 h after discontinuation, with follow-up to 24 h. Estimates are shown to three decimal places when a confidence limit rounds to 1.00. CI, confidence interval; HR, hazard ratio; IV, intravenous.*

**Table S10 Assessment of proportional hazards and interval-specific estimates**

*A. Tests of the proportional hazards assumption, P values*

| **Variable** | **Rebound hyperglycemia** | **Hypoglycemia** |
| --- | --- | --- |
| Glargine-to-IV dose ratio (per 0.1) | 0.020 | 0.52 |
| Glargine ≥2 h before discontinuation | 0.66 | 0.64 |
| Oral diet at discontinuation | 0.081 | 0.35 |

*B. Interval-specific adjusted hazard ratios (95% CI) for rebound hyperglycemia*

| **Interval after discontinuation** | **Episodes at risk** | **Events** | **Ratio, per 0.1 increase** | **Ratio ≥0.60 vs 0.40–0.49** | **Glargine ≥2 h before discontinuation** | **Oral diet at discontinuation** |
| --- | --- | --- | --- | --- | --- | --- |
| 0–6 h | 6,220 | 1,214 | 0.99 (0.95–1.02) | 0.94 (0.74–1.19) | 0.79 (0.69–0.92) | 1.53 (1.28–1.83) |
| 6–12 h | 4,411 | 500 | 0.930 (0.864–1.001) | 0.79 (0.55–1.15) | — | — |
| 12–24 h | 2,413 | 395 | 0.87 (0.81–0.94) | 0.63 (0.41–0.98) | — | — |
| 6–24 h | 4,411 | 895 | 0.91 (0.86–0.96) | — | 0.78 (0.67–0.91) | 1.42 (1.15–1.75) |

*Proportional hazards were assessed with score tests based on scaled Schoenfeld residuals in the fully adjusted models; the median P value across 20 imputations is shown. Interval-specific hazard ratios were estimated among episodes at risk at the start of each interval, with follow-up to the end of the interval, from fully adjusted models pooled over 20 imputations; dashes indicate estimates not calculated. Estimates are shown to three decimal places when a confidence limit rounds to 1.00. CI, confidence interval.*

**Table S11 Timing of the first glargine dose and the primary outcomes**

| **Timing of first glargine dose** | **Episodes, n** | **Rebound events** | **Rebound hyperglycemia within 24 h, HR (95% CI)** | **Rebound hyperglycemia within 6 h, HR (95% CI)** | **Hypoglycemia events** | **Hypoglycemia within 48 h, HR (95% CI)** |
| --- | --- | --- | --- | --- | --- | --- |
| After discontinuation | 355 | 166 | 1.05 (0.87–1.28) | 0.82 (0.64–1.06) | 10 | 0.74 (0.37–1.46) |
| 0 to <1 h before | 868 | 344 | 0.97 (0.85–1.11) | 0.77 (0.64–0.93) | 32 | 0.92 (0.59–1.43) |
| 1 to <2 h before (reference) | 3,107 | 939 | 1.00 | 1.00 | 87 | 1.00 |
| 2 to <3 h before | 1,559 | 452 | 0.87 (0.77–0.98) | 0.89 (0.76–1.04) | 49 | 0.97 (0.68–1.37) |
| ≥3 h before | 331 | 208 | 0.82 (0.67–1.01) | 0.60 (0.45–0.78) | 33 | 1.53 (0.90–2.61) |
| ≥2 h vs 0 to <2 h before (doses after discontinuation excluded) |  |  | 0.86 (0.77–0.96) | 0.867 (0.754–0.999) |  | 1.14 (0.85–1.53) |

*Hazard ratios are from Cox models adjusted for characteristics preceding the dose (all covariates except the infusion rate over the final 6 h, rate trend, final infusion rate and glucose values in the final 6 h, plus the mean infusion rate 6–12 h before discontinuation and the glargine dose per kilogram), pooled over 20 imputations. Of the 1,890 doses given at least 2 h before discontinuation, 575 were given 2.00–2.05 h before and 331 at least 3 h before. Estimates are shown to three decimal places when a confidence limit rounds to 1.00. CI, confidence interval; HR, hazard ratio.*

**Table S12 Glycemic outcomes and insulin use by nutritional status at discontinuation**

| **Variable** | **Nothing by mouth** | **Liquids or sips** | **Oral diet** | **Enteral tube feeding** | **Other or not recorded** |
| --- | --- | --- | --- | --- | --- |
| Episodes, n | 2,447 | 2,723 | 738 | 249 | 63 |
| Glargine-to-IV dose ratio | 0.40 (0.29–0.44) | 0.42 (0.33–0.46) | 0.41 (0.29–0.50) | 0.34 (0.20–0.46) | 0.33 (0.20–0.42) |
| Rebound hyperglycemia, events | 819 | 678 | 379 | 214 | 19 |
| Rebound hyperglycemia, cumulative incidence at 24 h, % (95% CI) | 41.8 (39.5–44.1) | 35.9 (33.5–38.4) | 64.8 (60.4–69.2) | 88.1 (83.6–91.9) | 40.9 (27.0–58.5) |
| Hypoglycemia, events | 85 | 67 | 35 | 22 | 2 |
| Hypoglycemia, cumulative incidence at 48 h, % (95% CI) | 5.8 (4.6–7.2) | 5.7 (4.2–7.7) | 8.4 (5.8–12.0) | 11.0 (7.4–16.4) | 3.2 (0.8–12.1) |
| Short-acting insulin within 24 h, n (%) | 1,977 (80.8) | 2,062 (75.7) | 644 (87.3) | 235 (94.4) | 49 (77.8) |
| Short-acting insulin dose within 24 h among recipients, U | 8 (4–16) | 6 (4–13) | 12 (6–24) | 26 (16–50) | 8 (4–14) |
| Short-acting insulin doses within 24 h among recipients, n | 2 (1–3) | 2 (1–3) | 2 (1–4) | 4 (3–5) | 2 (1–3) |
| Mean glucose within 24 h, mg/dL | 136 (120–159) | 133 (119–153) | 153 (129–189) | 193 (159–241) | 135 (124–171) |

*Nutritional status was the most recent “Diet Type” entry within 24 h before discontinuation; “other or not recorded” includes 11 episodes with parenteral nutrition. Values are median (interquartile range) unless stated otherwise. Cumulative incidence was estimated with the Kaplan–Meier method with censoring at ICU discharge or resumption of intravenous insulin. Short-acting insulin comprised non-infusion doses of rapid-acting analogues and regular insulin given after discontinuation; mean glucose refers to episodes with at least one glucose value during the first 24 h. CI, confidence interval; ICU, intensive care unit.*

**Table S13 Association between the glargine-to-intravenous dose ratio and the primary outcomes in predefined subgroups**

*A. Entire follow-up*

| **Subgroup** | **Level** | **Episodes, n** | **Rebound events** | **Rebound HR per 0.1 (95% CI)** | **P for interaction** | **Hypoglycemia events** | **Hypoglycemia HR per 0.1 (95% CI)** | **P for interaction** |
| --- | --- | --- | --- | --- | --- | --- | --- | --- |
| Diabetes | No | 3,359 | 424 | 0.97 (0.91–1.03) | 0.69 | 75 | 1.08 (0.96–1.21) | 0.55 |
|  | Yes | 2,861 | 1,685 | 0.96 (0.93–0.99) |  | 136 | 1.12 (1.05–1.19) |  |
| HbA1c ≥8.0% (observed values) | No | 5,034 | 1,288 | 0.94 (0.90–0.98) | 0.28 | 144 | 1.08 (0.98–1.19) | 0.38 |
|  | Yes | 784 | 553 | 0.97 (0.93–1.02) |  | 40 | 1.14 (1.05–1.23) |  |
| Insulin before admission | No | 4,285 | 1,041 | 0.93 (0.89–0.98) | 0.19 | 111 | 1.12 (1.01–1.24) | 0.59 |
|  | Yes | 1,053 | 726 | 0.966 (0.931–1.004) |  | 78 | 1.08 (0.99–1.19) |  |
| Cardiac surgery | No | 780 | 494 | 0.96 (0.92–1.01) | 0.81 | 49 | 1.111 (0.996–1.239) | 0.99 |
|  | Yes | 5,440 | 1,615 | 0.96 (0.92–0.99) |  | 162 | 1.11 (1.04–1.19) |  |
| Oral diet at discontinuation | No | 5,482 | 1,730 | 0.94 (0.90–0.97) | 0.011 | 176 | 1.09 (1.01–1.17) | 0.36 |
|  | Yes | 738 | 379 | 1.00 (0.96–1.04) |  | 35 | 1.14 (1.05–1.24) |  |
| Glargine ≥2 h before discontinuation | No | 4,330 | 1,449 | 0.94 (0.91–0.98) | 0.068 | 129 | 1.16 (1.08–1.24) | 0.062 |
|  | Yes | 1,890 | 660 | 0.98 (0.95–1.02) |  | 82 | 1.04 (0.93–1.16) |  |
| Corticosteroid therapy | No | 5,990 | 1,936 | 0.95 (0.92–0.98) | 0.25 | 194 | 1.11 (1.04–1.18) | 0.32 |
|  | Yes | 230 | 173 | 1.00 (0.92–1.09) |  | 17 | 1.220 (1.002–1.485) |  |
| Age ≥65 years | No | 2,468 | 831 | 0.95 (0.91–0.99) | 0.49 | 73 | 1.11 (1.01–1.21) | 0.88 |
|  | Yes | 3,752 | 1,278 | 0.966 (0.930–1.003) |  | 138 | 1.12 (1.04–1.20) |  |
| Sex | Male | 4,409 | 1,447 | 0.965 (0.931–1.000) | 0.54 | 128 | 1.13 (1.03–1.23) | 0.69 |
|  | Female | 1,811 | 662 | 0.95 (0.91–0.99) |  | 83 | 1.10 (1.02–1.19) |  |

*B. Oral diet at discontinuation: rebound hyperglycemia within 6 h and between 6 and 24 h*

| **Interval** | **Oral diet** | **Episodes at risk** | **Events** | **Rebound hyperglycemia, HR per 0.1 (95% CI)** | **P for interaction** |
| --- | --- | --- | --- | --- | --- |
| 0–6 h | No | 5,482 | 962 | 0.956 (0.914–1.000) | 0.003 |
|  | Yes | 738 | 252 | 1.04 (0.99–1.09) |  |
| 6–24 h | No | 4,011 | 768 | 0.91 (0.86–0.97) | 0.88 |
|  | Yes | 400 | 127 | 0.90 (0.83–0.98) |  |

*Hazard ratios per 0.1 increase in the ratio were estimated from fully adjusted Cox models including a product term between the ratio and the subgroup indicator, pooled over 20 imputations. The HbA1c subgroup analysis was restricted to episodes with a measured value and the analysis by insulin use before admission to episodes with known status. In panel B, the analysis for 0–6 h includes all episodes and that for 6–24 h includes episodes still at risk of rebound hyperglycemia 6 h after discontinuation. Estimates are shown to three decimal places when a confidence limit rounds to 1.00. CI, confidence interval; HbA1c, glycated hemoglobin; HR, hazard ratio.*

**Table S14 Sensitivity analyses**

| **Analysis** | **Episodes, n** | **Rebound events** | **Rebound, per 0.1 increase** | **Rebound, ≥0.60 vs 0.40–0.49** | **Hypoglycemia events** | **Hypoglycemia, per 0.1 increase** | **Hypoglycemia, ≥0.60 vs 0.40–0.49** |
| --- | --- | --- | --- | --- | --- | --- | --- |
| Primary analysis | 6,220 | 2,109 | 0.96 (0.93–0.99) | 0.85 (0.70–1.02) | 211 | 1.11 (1.04–1.18) | 1.76 (1.09–2.83) |
| (1) Complete-case analysis | 5,790 | 1,834 | 0.96 (0.92–0.99) | 0.85 (0.70–1.04) | 181 | 1.11 (1.03–1.19) | 1.78 (1.06–3.00) |
| (2) Inverse probability of censoring weighting | 6,220 | 2,109 | 0.96 (0.93–0.99) | 0.85 (0.69–1.03) | 211 | 1.13 (1.06–1.20) | 1.68 (1.03–2.74) |
| (3) Potential ICU follow-up of at least 24 h | 2,742 | 1,208 | 0.95 (0.91–0.99) | 0.80 (0.62–1.02) | 155 | 1.11 (1.01–1.22) | 1.39 (0.77–2.50) |
| (4) IV total daily dose from the mean rate over up to 24 h | 6,220 | 2,109 | 0.93 (0.90–0.96) | 0.773 (0.598–1.000) | 211 | 1.10 (1.01–1.20) | 2.42 (1.35–4.34) |
| (5) First eligible episode of each patient | 5,924 | 1,892 | 0.95 (0.92–0.98) | 0.82 (0.67–1.01) | 178 | 1.10 (1.02–1.19) | 1.59 (0.94–2.68) |
| (6) Excluding glargine given after discontinuation | 5,865 | 1,943 | 0.96 (0.93–0.99) | 0.84 (0.70–1.02) | 201 | 1.11 (1.04–1.19) | 1.77 (1.09–2.89) |
| (7) Excluding point-of-care glucose <70 mg/dL in the final 6 h | 6,118 | 2,070 | 0.96 (0.93–0.99) | 0.86 (0.71–1.04) | 202 | 1.11 (1.04–1.18) | 1.76 (1.08–2.89) |
| (8) Excluding basal insulin 12–48 h before discontinuation | 5,865 | 1,834 | 0.95 (0.91–0.99) | 0.82 (0.66–1.01) | 173 | 1.18 (1.09–1.27) | 1.71 (1.01–2.90) |
| (9) Exposure from ICU charting alone | 6,138 | 2,093 | 0.96 (0.93–0.99) | 0.84 (0.70–1.02) | 210 | 1.11 (1.04–1.18) | 1.79 (1.11–2.90) |
| (10) All basal insulin types | 6,441 | 2,294 | 0.96 (0.93–0.99) | 0.86 (0.71–1.03) | 230 | 1.12 (1.05–1.19) | 1.83 (1.16–2.88) |
| (11) Excluding type 1 diabetes | 6,046 | 1,978 | 0.95 (0.92–0.99) | 0.80 (0.66–0.98) | 180 | 1.11 (1.03–1.19) | 1.89 (1.14–3.15) |
| (12) Parsimonious adjustment set | 6,220 | 2,109 | 0.95 (0.93–0.98) | 0.86 (0.72–1.03) | 211 | 1.15 (1.09–1.21) | 2.07 (1.31–3.27) |
| (13) Linear terms for continuous covariates | 6,220 | 2,109 | 0.98 (0.95–1.01) | 0.95 (0.79–1.15) | 211 | 1.10 (1.03–1.17) | 1.66 (1.04–2.67) |
| (14) Excluding ratios of 10/24 (0.417) | 5,172 | 1,892 | 0.96 (0.93–0.99) | 0.84 (0.69–1.03) | 187 | 1.11 (1.04–1.18) | 1.58 (0.92–2.71) |
| (15) Ratios of 1.0 or less | 6,145 | 2,074 | 0.94 (0.90–0.97) | 0.84 (0.69–1.02) | 207 | 1.21 (1.09–1.34) | 1.92 (1.18–3.12) |
| (16) Excluding the 2020–2022 anchor year group | 5,809 | 1,960 | 0.96 (0.93–0.99) | 0.85 (0.70–1.03) | 207 | 1.11 (1.04–1.18) | 1.68 (1.04–2.71) |
| (17) Hypoglycemia within 24 h | 6,220 | — | — | — | 155 | 1.12 (1.05–1.20) | 2.17 (1.27–3.70) |
| (18) Hypoglycemia events preceded by short-acting insulin censored | 6,220 | — | — | — | 179 | 1.09 (1.02–1.17) | 1.59 (0.95–2.68) |
| (19) Ratio and infusion rate recalculated over running time | 6,220 | 2,109 | 0.964 (0.934–0.996) | 0.87 (0.70–1.09) | 211 | 1.10 (1.03–1.18) | 1.56 (0.92–2.65) |
| (20) Excluding interruptions ≥15 min in the final 6 h | 5,291 | 1,813 | 0.96 (0.92–0.99) | 0.85 (0.67–1.08) | 177 | 1.10 (1.03–1.19) | 1.69 (0.93–3.07) |
| (21) Excluding interruptions ≥30 min after the glargine dose | 6,183 | 2,090 | 0.96 (0.93–0.99) | 0.85 (0.70–1.03) | 211 | 1.11 (1.05–1.19) | 1.86 (1.15–3.00) |
| Alternative exposure: glargine dose per 0.1 U/kg | 6,220 | 2,109 | 0.93 (0.90–0.98) | — | 211 | 1.17 (1.04–1.30) | — |

*Values are adjusted hazard ratios (95% confidence intervals) pooled over 20 imputations, except for analysis (1), which used complete cases, and analysis (2), which reports odds ratios from weighted pooled logistic models. For the weight-based exposure, hazard ratios per 0.1 U/kg were also estimated for severe hyperglycemia (HR 0.90, 0.84–0.97) and resumption of intravenous insulin (HR 0.86, 0.79–0.94). The E-value for the association between ratios of 0.60 or higher and hypoglycemia was 2.92 (1.41 for the lower confidence limit), and that for each 0.1 increase in the ratio was 1.46 (1.26). Estimates are shown to three decimal places when a confidence limit rounds to 1.00. In analysis 19, 637 episodes changed ratio category and 286 had a ratio of 0.60 or higher. An interruption of 15 min or longer within the final 6 h occurred in 929 episodes (14.9%): 170 (8.1%) in the 0.40–0.49 category, 169 (30.8%) in the 0.50–0.59 category and 324 (57.1%) at ratios of 0.60 or higher; interruptions of 30 min or longer after the glargine dose occurred in 37 episodes. ICU, intensive care unit; IV, intravenous.*

**Table S15 Glycemic control and insulin use after discontinuation by glargine-to-intravenous dose ratio**

| **Variable** | **Overall** | **<0.30** | **0.30–0.39** | **0.40–0.49 (reference)** | **0.50–0.59** | **≥0.60** |
| --- | --- | --- | --- | --- | --- | --- |
| Episodes with ≥1 glucose value, n (%) | 6,075 (97.7) | 1,503 (98.4) | 1,438 (97.6) | 2,049 (97.4) | 531 (96.9) | 554 (97.7) |
| Glucose values per episode | 4 (2–6) | 5 (3–6) | 4 (2–6) | 4 (2–6) | 4 (2–6) | 5 (3–6) |
| ICU follow-up within 24 h, h | 15.2 (8.9–24.0) | 20.5 (9.8–24.0) | 14.6 (8.7–24.0) | 14.2 (8.7–24.0) | 14.8 (8.6–24.0) | 17.8 (9.4–24.0) |
| Glucose values per hour of follow-up | 0.25 (0.21–0.32) | 0.26 (0.21–0.34) | 0.25 (0.21–0.31) | 0.25 (0.21–0.29) | 0.25 (0.21–0.33) | 0.26 (0.21–0.33) |
| Mean glucose, mg/dL | 138 (121–163) | 144 (126–178) | 138 (122–162) | 133 (118–154) | 136 (119–161) | 137 (121–165) |
| Values within 70–180 mg/dL, mean % | 81.5 | 74.8 | 81.7 | 86.6 | 83.1 | 78.7 |
| Values >180 mg/dL, mean % | 18.0 | 24.7 | 18.0 | 12.9 | 16.2 | 19.8 |
| Glucose standard deviation, mg/dL | 22.6 (14.4–35.6) | 25.6 (16.5–43.2) | 22.4 (14.0–33.9) | 20.0 (12.9–31.2) | 22.2 (14.7–36.0) | 25.4 (16.2–39.4) |
| Maximum glucose, mg/dL | 160 (136–200) | 172 (143–227) | 159 (136–197) | 154 (132–183) | 157 (136–194) | 167 (138–204) |
| Short-acting insulin within 24 h, n (%) | 4,967 (79.9) | 1,291 (84.5) | 1,192 (80.9) | 1,596 (75.9) | 424 (77.4) | 464 (81.8) |
| Short-acting insulin dose within 24 h among recipients, U | 8 (4–16) | 10 (4–21) | 8 (4–16) | 8 (4–14) | 8 (4–14) | 8 (4–18) |
| Time to first short-acting dose among recipients, h | 4.3 (1.9–6.1) | 3.5 (1.6–5.9) | 4.5 (1.9–6.2) | 4.7 (2.1–6.2) | 4.6 (1.9–6.0) | 3.7 (1.6–6.0) |
| Additional basal insulin within 48 h, n (%) | 1,453 (23.4) | 463 (30.3) | 325 (22.1) | 376 (17.9) | 117 (21.4) | 172 (30.3) |
| Hypoglycemia within 48 h preceded by short-acting insulin within 4 h, n/N (%) | 32/211 (15.2) | 8/57 (14.0) | 7/36 (19.4) | 4/58 (6.9) | 4/22 (18.2) | 9/38 (23.7) |

*Values are median (interquartile range), number (%) or mean percentage, as indicated. Glucose metrics refer to ICU follow-up within 24 h after discontinuation (until ICU discharge or resumption of intravenous insulin, if earlier), and glucose values per hour were calculated over this follow-up; percentages of values are averaged across episodes with at least one glucose value, and the standard deviation was calculated for episodes with at least three values (n = 4,439). Short-acting insulin comprised non-infusion doses of rapid-acting analogues and regular insulin charted in the ICU or recorded in the electronic medication administration record (eMAR); regular insulin boluses could not be distinguished between subcutaneous and intravenous administration. Excluding episodes with any short-acting or basal insulin record derived only from the eMAR (n = 1,215), short-acting insulin was given in 75.5% of episodes. Additional basal insulin refers to basal doses given more than 1 h after discontinuation during the 48-h follow-up. eMAR, electronic medication administration record; ICU, intensive care unit.*
